# Poor *in-utero* growth, and reduced beta cell compensation and high fasting glucose from childhood, are harbingers of glucose intolerance in young Indians

**DOI:** 10.1101/2020.11.19.20234542

**Authors:** Chittaranjan S Yajnik, Souvik Bandopadhyay, Aboli Bhalerao, Dattatray S Bhat, Sanat B Phatak, Rucha H Wagh, Pallavi C Yajnik, Anand Pandit, Sheila Bhave, Kurus Coyaji, Kalyanaraman Kumaran, Clive Osmond, Caroline HD Fall

## Abstract

**Objective:** India is a double world capital for early life undernutrition and type 2 diabetes. We aimed to characterise lifecourse growth and metabolic trajectories in those developing glucose intolerance as young adults, in the Pune Maternal Nutrition Study (PMNS).

**Research design and Methods:** PMNS is a community-based intergenerational birth cohort established in 1993, with serial information on parents and children through pregnancy, childhood and adolescence. We compared normal glucose tolerant and glucose intolerant participants for serial growth, estimates of insulin sensitivity and secretion (HOMA and dynamic indices) and beta cell compensation accounting for prevailing insulin sensitivity (disposition index).

**Results:** At 18 years (N=619) 37% men and 20% women were glucose intolerant (184 prediabetes, 1 diabetes) despite 48% being underweight (BMI<18.5 kg/m^2^). Glucose intolerant participants had higher fasting glucose from childhood. Mothers of glucose intolerant participants had higher glycemia in pregnancy. Glucose intolerant participants were shorter at birth. Insulin sensitivity decreased with age in all participants, and the glucose intolerant had consistently lower compensatory insulin secretion from childhood. Participants in the highest quintile of fasting glucose at 6 and 12 years had a 2.5- and 4.0-fold higher risk respectively of 18-year glucose intolerance; this finding was replicated in two other cohorts.

**Conclusion:** Inadequate compensatory insulin secretory response to increasing insulin insensitivity from early life is the major pathophysiology underlying glucose intolerance in thin rural Indians. Smaller birth size, maternal pregnancy hyperglycemia, and higher glycemia in childhood herald future glucose intolerance, mandating a strategy for diabetes prevention from early life, preferably intergenerationally.

## Introduction

India is experiencing a rapidly escalating epidemic of type 2 diabetes, (1) and simultaneously has the world’s highest burden of low birthweight and under-five undernutrition. (2) Current thinking about the etiology of type 2 diabetes is mostly based on studies in adults and ascribes it to overnutrition and sedentariness in genetically susceptible individuals. On this background, the high prevalence of diabetes in Indians, at a younger age and lower body mass index (BMI) than Europeans, appears paradoxical. (3) Recent reports suggest high rates of prediabetes in Indian adolescents and young adults (2) and faster conversion from prediabetes to diabetes. (4,5) The greatest rise in prevalence in the last 25 years has occured in the most deprived Indian states, and in some places there is a reversal of socioeconomic trends from a previous excess prevalence among the most affluent. (6) Taken together, these findings raise the possibility that historical deprivation and undernutrition are contributory factors to diabetes in a rapidly transitioning society like India.

There is growing acceptance of a lifecourse model (Developmental Origins of Health and Disease, DOHaD) for the evolution of type 2 diabetes. Adverse environmental exposures in early life, classically reflected in low birth weight, are associated with an increased risk of adult type 2 diabetes. (7,8) The ‘thrifty phenotype’ hypothesis proposed that intra-uterine undernutrition disrupts the structure and function of key organs, which manifests as an increased risk of diabetes through both diminished insulin secretion and sensitivity. (9) While there is considerable information on newborn size and childhood growth as predictors of later type 2 diabetes risk, (10,11) there is little data on childhood measures of glucose, insulin secretion and sensitivity as predictors. It is therefore unknown at what age metabolic susceptibility to future diabetes becomes evident, and whether impaired insulin insensitivity or secretion is the primary defect. Consequently, diabetes prevention trials still mainly target middle aged people who already have obesity and advanced metabolic abnormalities. (12)

In the Pune Maternal Nutrition Study (PMNS), we had a unique opportunity to construct the first ever lifecourse trajectory of glucose-insulin indices and growth in young rural Indian adults, along with data on parental size and glucose intolerance.

## Methods

### Overview of the PMNS cohort

The PMNS (Fig. 1, Supplemental Fig. S1) was established in 1993 in six villages near Pune, India to prospectively study associations of maternal nutritional status with fetal growth and later diabetes risk in the offspring. (13) Married, non-pregnant women (F0 generation, N=2,466) were followed up and those who became pregnant (N=797) were recruited into the study if a singleton pregnancy of <21 weeks’ gestation was confirmed by ultrasound. Most delivered at home and only 4.2% required Caesarean section; 3 women had diabetes in pregnancy (WHO criteria, 1985).

**Figure 1:**
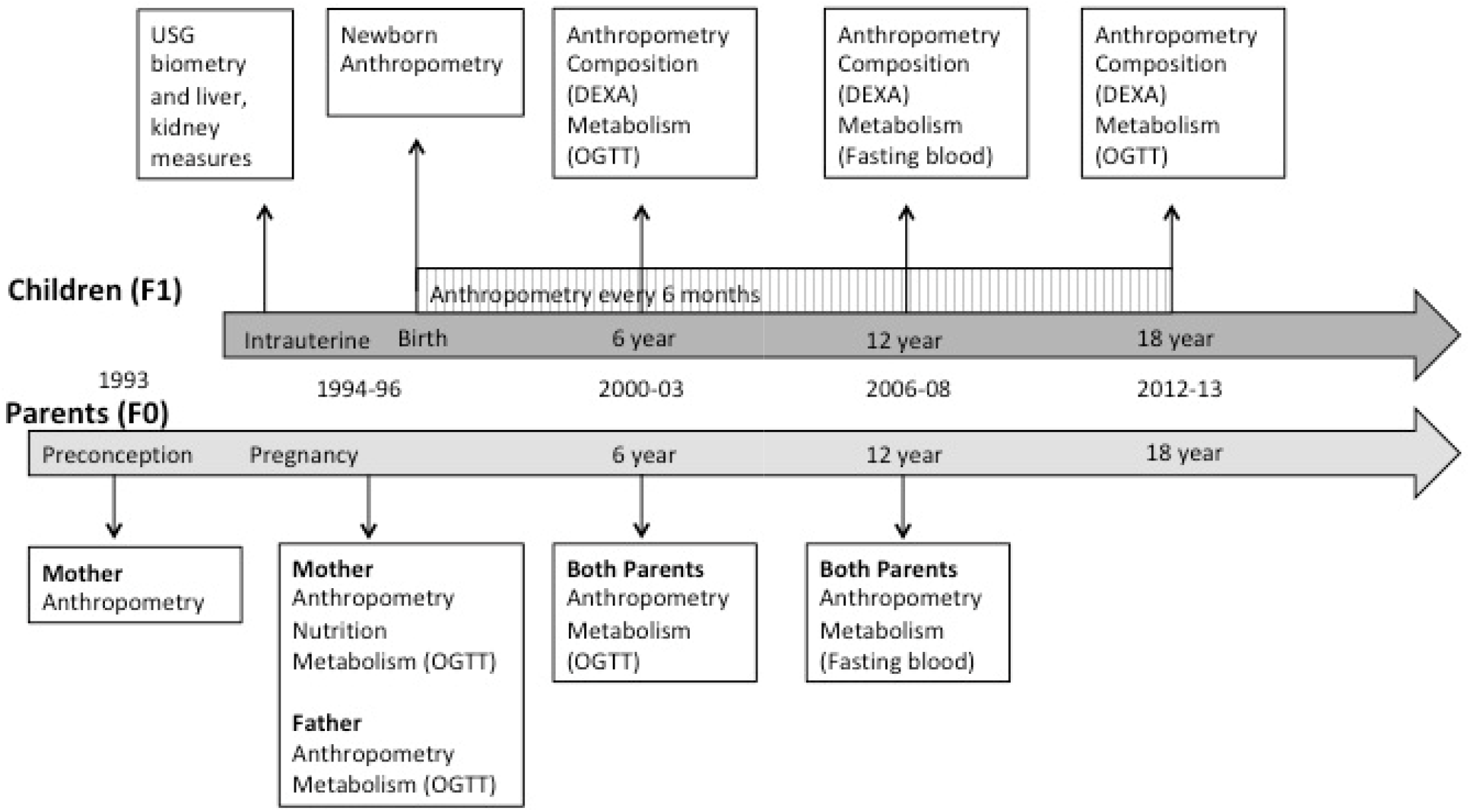
The Pune Maternal Nutrition Study. Married, non-pregnant women were followed up in six villages near Pune, India. Those who became pregnant (singleton fetus, less than 21 weeks) were enrolled during pregnancy. Maternal glucose tolerance was measured at 28 ±2 weeks’ gestation. Babies (F1 generation) were measured at birth and every 6 months thereafter by detailed anthropometry. Comprehensive measurements of body size and composition, and glucose-insulin metabolic function were performed every six years in the children until age 18 years and in both parents (F0 generation) when the child was aged 6 and 12 years. USG: Ultrasonography; DEXA: Dual Energy X-ray Absorptiometry; OGTT: Oral Glucose Tolerance Test.

### Measurements of babies and children (F1 generation)

Detailed anthropometry was carried out using standardized methods at birth and every 6 months post-natally. (14) Glucose and insulin concentrations, body composition and socio-economic status (SES) were measured at age 6, 12 and 18 years at the Diabetes Unit. All families were visited by field staff a week before the study to explain the procedures and to stress that they should eat normally and perform usual daily activities. Participants arrived at the Diabetes Unit the evening before the investigations, had a standard dinner, and fasted overnight. In the morning, a fasting blood sample was collected. At age 6 years, an oral glucose tolerance test (OGTT, 1.75g/kg anhydrous glucose) was performed. At 12 years, only a fasting sample was collected. At 18 years an OGTT (75g anhydrous glucose) was repeated. Glucose was measured by the glucose oxidase/peroxidase method, and specific insulin by ELISA (CV for glucose <4%, insulin <8% at all time points). Insulin assays were calibrated against the same WHO standard (WHO 1^st^ IRP (66/304) and are therefore directly comparable (Supplemental Table S3). Insulin sensitivity (HOMA-S) and beta cell function (HOMA-β) were calculated using data from the fasting samples on https://www.phc.ox.ac.uk/research/technology-outputs/ihoma2, last accessed August 2019. We calculated the Matsuda Index for insulin sensitivity (15, 16) and the Insulinogenic Index (ln{Insulin(30-min/fasting) /Glucose(30-min/fasting)} for early insulin secretion. (17) Both indices are validated against reference methods and used commonly in epidemiological research. Because of the dependence of the beta cell response on prevailing insulin sensitivity, we also calculated the Disposition index (insulinogenic index*Matsuda index) at 6 and 18 years and HOMA-S*HOMA-β at 12 years to estimate beta cell compensatory response. (18) Total fat and lean mass and body fat% were measured by Dual Energy X-ray Absorptiometry (DXA). SES was evaluated using the Standard of Living Index (SLI) based on the family’s dwelling, land ownership and assets. (19) Higher scores denote higher SES.

### Definitions

In adults, underweight was defined as a BMI <18.5 kg/m^2^, overweight/obesity as a BMI ≥25 kg/m^2^ (WHO international cut point) and stunting as a height-for-age Z-score <-2 SD below the WHO average (<149.8 cm in women and <161.2 cm for men; [http://www.who.int/growthref/who2007_height_for_age/en/, last accessed August 2020]). Central obesity was defined as a waist circumference >90 cm (men) and >80 cm (women) [https://www.idf.org/e-library/consensus-statements/60-idfconsensus-worldwide-definitionof-the-metabolic-syndrome.html, last accessed August 2020] and adiposity as a DXA-derived fat% >25% (men) and >35% (women). Glucose tolerance in children, fathers and non-pregnant mothers was classified by ADA criteria (20) as normal (NGT), prediabetes (impaired fasting glucose [IFG] or impaired glucose tolerance [IGT]), or diabetes mellitus. IFG, IGT and diabetes together were referred to as ‘glucose intolerance’.

### Parental measurements

Anthropometry and glucose tolerance (75g OGTT) were measured in both parents during the index pregnancy (∼28 weeks gestation) and at the 6-year follow up. Gestational diabetes was diagnosed by WHO 1985 criteria [2-h plasma glucose ≥7.8 mmol/l] and treated appropriately. Given the small number of GDM cases, for the current analysis we defined glucose intolerance as fasting plasma glucose (FPG) ≥95^th^ centile in this population (5.1 mmol/l), which coincides with current IADPSG criteria. (21) Anthropometry and only a fasting blood test were available at the 12-year follow-up. Parents were classified as ever underweight or overweight/obese based on their follow-up data. Fathers and mothers were classified as glucose intolerant if they had IFG, IGT or diabetes at any follow up.

### Replication cohorts

#### Pune Children’s Study Cohort (PCS)

PCS is an urban cohort of children born in the KEM Hospital in 1987-89. (22) Briefly, the children were studied at 8-years (n=477) and 21 years (n=357) of age. Measurements were the same as those in the PMNS, and glucose tolerance at age 21 years was classified by the same ADA criteria.

#### Extended PMNS cohort

This cohort included an additional 110 pregnancies after completing recruitment of the main PMNS cohort, to validate ultrasound protocols for gestational dating. Ninety-two children had glucose tolerance data at 6, 12 and 18 years of age. Given the small numbers in this cohort, we used the upper tertile of 18-year FPG concentration as the outcome.

#### Statistical methods

Our purpose was to show a comparative temporal evolution of glucose-insulin relationships and growth in prediabetic and normal glucose tolerant young adults at 18 years (Table 1). Variables with right-skewed distributions were log transformed; all variables were Z-standardised, and differences between NGT and glucose intolerant participants were expressed in Z-score units with 95% confidence intervals. We used logistic regression for lifecourse predictors of glucose intolerance at 18 years of age (outcome). The predictors included parental body size and glucose tolerance; the F1 participants’ birth measurements and childhood and adolescent body size and glucose concentrations, in addition to sex and SES. Thus, our analysis includes a combination of traditional and novel risk factors representing the DOHaD paradigm. We used interaction tests to investigate whether associations differed between the sexes. We created ROC curves to show the sensitivity and specificity of these variables in predicting glucose intolerance.

**Table 1:** Comparison of biomarkers between participants with normal glucose tolerance and glucose intolerant at age 18 years.

|  | MEN |  |  |  |  | WOMEN |  |  |  |  | SEXES COMBINED |  |  |
| --- | --- | --- | --- | --- | --- | --- | --- | --- | --- | --- | --- | --- | --- |
|  | NGT<br>(median)<br>N=221 | Glucose<br>intolerant<br>(median)<br>N=131 | Mean<br>Difference<br>(Z-score)<br>Glucose<br>intolerant<br>– NGT, | 95% CI of<br>Mean<br>Difference | P | NGT<br>(median)<br>N=213 | Glucose<br>intolerant<br>(median)<br>N=54 | Mean<br>Difference<br>(Z-score)<br>Glucose<br>intolerant<br>– NGT, | 95% CI of<br>Mean<br>Difference | P | Mean<br>Difference<br>(Z-score)<br>Glucose<br>intolerant<br>– NGT | 95% CI<br>of Mean<br>Difference | P |
| **18 YEARS** |  |  |  |  |  |  |  |  |  |  |  |  |  |
| Height (cm) | **170.1** | **169.0** | **-0.19** | (-0.40,<br>0.03) | 0.09 | **157.2** | **157.3** | **-0.14** | (-0.44, 0.16) | 0.35 | **-0.17** | (-<br>0.35,0.00) | 0.06 |
| BMI (kg/m<sup>2</sup>) | **18.8** | **19.6** | **0.29** | (0.07, 0.50) | 0.009 | **18.1** | **17.4** | **-0.18** | (-0.48, 0.12) | 0.23 | **0.12** | (-0.05,<br>0.30) | 0.16 |
| Waist circumference (cm) | **70.5** | **72.6** | **0.30** | (0.09, 0.52) | 0.006 | **67.4** | **66.3** | **-0.2** | (-0.50, 0.10) | 0.19 | **0.13** | (-0.04,<br>0.31) | 0.14 |
| Fat percentage (%) | **12.5** | **14.8** | **0.37** | (0.16, 0.59) | <0.001 | **28.1** | **27.0** | **-0.03** | (-0.33, 0.28) | 0.87 | **0.24** | (0.06,<br>0.41) | 0.008 |
| SES (SLI score) | **37** | **38** | **0.20** | (-0.02,<br>0.41) | 0.07 | **36** | **36** | **-0.09** | (-0.39, 0.21) | 0.55 | **0.10** | (-0.08,<br>0.27) | 0.27 |
| Fasting glucose (mmol/l) | **5.2** | **5.7** | **1.49** | (1.35, 1.64) | <0.001 | **5.1** | **5.6** | **1.19** | (0.93, 1.45) | <0.001 | **1.39** | (1.25,<br>1.52) | <0.001 |
| 30-min glucose (mmol/l) | **7.8** | **8.9** | **0.81** | (0.62, 1.01) | <0.001 | **8.0** | **9.1** | **0.95** | (0.68, 1.23) | <0.001 | **0.86** | (0.68,<br>0.99) | <0.001 |
| 120-min glucose (mmol/l) | **5.7** | **7.0** | **0.92** | (0.73, 1.12) | <0.00<br>1 | **6.0** | **7.9** | **1.39** | (1.14, 1.63) | <0.00<br>1 | **1.08** | (0.93, 1.23) | <0.00<br>1 |
| Fasting insulin (pmol/l) | **48.0** | **66.0** | **0.57** | (0.36, 0.78) | <0.00<br>1 | **63.0** | **66.6** | **0.22** | (-0.08, 0.53) | 0.15 | **0.45** | (0.28, 0.63) | <0.00<br>1 |
| 30-min insulin (pmol/l) | **477.0** | **477.6** | **0.07** | (-0.13, 0.31) | 0.54 | **578.4** | **588.3** | **-0.16** | (-0.52, 0.09) | 0.30 | **-0.01** | (-0.19, 0.16) | 0.92 |
| 120-min insulin (pmol/l) | **258.0** | **352.2** | **0.51** | (0.30, 0.72) | <0.00<br>1 | **360.0** | **579.6** | **0.84** | (0.56, 1.13) | <0.00<br>1 | **0.62** | (0.43, 0.77) | <0.00<br>1 |
| HOMA-S | **97** | **69** | **-0.63** | (-0.83, -0.42) | <0.00<br>1 | **75** | **70** | **-0.26** | (-0.57, 0.04) | 0.09 | **-0.51** | (-0.68, -0.33) | <0.00<br>1 |
| HOMA-β | **92** | **94** | **0.04** | (-0.18, 0.26) | 0.71 | **117** | **110** | **-0.28** | (-0.58, 0.03) | 0.07 | **-0.07** | (-0.24, 0.11) | 0.46 |
| Disposition Index (HOMA) | **84.2** | **64.3** | **-1.25** | (-1.39, -1.01) | <0.00<br>1 | **84.1** | **71.3** | **-0.98** | (-1.24, -0.68) | <0.00<br>1 | **-1.16** | (-1.23, -0.91) | <0.00<br>1 |
| Insulinogenic Index<sup>a</sup> | **1.83** | **1.53** | **-0.49** | (-0.70, -0.28) | <0.00<br>1 | **1.77** | **1.53** | **-0.56** | (-0.85, -0.26) | <0.00<br>1 | **-0.52** | (-0.69, -0.34) | <0.00<br>1 |
| Matsuda Index<sup>b</sup> | **15.4** | **11.3** | **-0.70** | (-0.90, -0.50) | <0.00<br>1 | **12.2** | **9.07** | **-0.69** | (-0.98, -0.48) | <0.00<br>1 | **-0.67** | (-0.81, -0.54) | <0.00<br>1 |
| Disposition Index<sup>c</sup> | **4.56** | **4.02** | **-0.55** | (-0.76, -0.34) | <0.00<br>1 | **4.26** | **3.66** | **-0.84** | (-1.13, -0.56) | <0.00<br>1 | **-0.63** | (-0.79, -0.46) | <0.00<br>1 |
| **15 YEARS** |  |  |  |  |  |  |  |  |  |  |  |  |  |
| HbA1c (%) | **5.40** | **5.50** | **0.21** | (-0.03, 0.44) | 0.082 | **5.30** | **5.40** | **0.18** | (-0.14, 0.49) | 0.269 | **0.19** | (0.01, 0.38) | 0.043 |
| HbA1c (mmol/mol) <sup>e</sup> | **35.51** | **36.61** | **0.21** | (-0.03, 0.44) | 0.082 | **34.42** | **35.51** | **0.18** | (-0.14, 0.49) | 0.269 | **0.19** | (0.01, 0.38) | 0.043 |
| **12 YEARS** |  |  |  |  |  |  |  |  |  |  |  |  |  |
| Height (cm) | **138.9** | **137.4** | **-0.20** | (-0.42, 0.02) | 0.07 | **139.6** | **139.1** | **-0.17** | (-0.47, 0.14) | 0.28 | **-0.19** | -0.36, -0.01 | 0.04 |
| BMI (kg/m<sup>2</sup>) | **14.6** | **14.7** | **0.02** | (-0.20, 0.23) | 0.88 | **14.4** | **14.2** | **-0.08** | (-0.39, 0.22) | 0.59 | **-0.02** | -0.19, 0.16 | 0.85 |
| Waist circumference (cm) | **57.2** | **57.2** | **0.02** | (-0.21, 0.24) | 0.89 | **56.0** | **55.0** | **-0.36** | (-0.67, -0.06) | 0.02 | **-0.11** | -0.29, 0.07 | 0.21 |
| Fat percentage (%) | **13.4** | **14.5** | **0.19** | (-0.04, 0.41) | 0.10 | **17.8** | **17.1** | **-0.02** | (-0.33, 0.3) | 0.92 | **0.12** | -0.06, 0.30 | 0.20 |
| Fasting glucose (mmol/l) | **4.8** | **4.9** | **0.40** | (0.19, 0.62) | <0.001 | **4.7** | **5.1** | **0.82** | (0.54, 1.11) | <0.001 | **0.55** | 0.38, 0.72 | <0.001 |
| Fasting insulin (pmol/l) | **27.6** | **30.6** | **0.10** | (-0.12, 0.32) | 0.37 | **34.0** | **34.5** | **-0.01** | (-0.31, 0.3) | 0.97 | **0.06** | -0.11, 0.24 | 0.48 |
| HOMA-S | **165** | **153** | **-0.12** | (-0.34, 0.10) | 0.28 | **138** | **134** | **-0.02** | (-0.32, 0.28) | 0.89 | **-0.09** | -0.26, 0.09 | 0.34 |
| HOMA-β | **71** | **73** | **-0.06** | (-0.28, 0.16) | 0.61 | **90** | **80** | **-0.39** | (-0.69, -0.09) | 0.01 | **-0.17** | -0.35, 0.01 | 0.06 |
| Disposition Index (HOMA)<sup>d</sup> | **123** | **112** | **-0.33** | (-0.54, -0.11) | 0.003 | **122** | **108** | **-0.58** | (-0.88, -0.29) | <0.001 | **-0.41** | -0.59, -0.24 | <0.001 |
| Pubertal stage <sup>s</sup> | **1** | **1** |  |  | 0.630 | **2** | **2** |  |  | 0.136 |  |  |  |
| Age at menarche (y) | - | - | - | - | - | **13.5** | **13.8** | **0.22** | (-0.11, 0.56) | 0.196 | - | - | - |
| **6 YEARS** |  |  |  |  |  |  |  |  |  |  |  |  |  |
| Height (cm) | **110.0** | **110.0** | **0.01** | (-0.21, 0.23) | 0.92 | **109.4** | **108.2** | **-0.38** | (-0.67, -0.08) | 0.01 | **-0.12** | (-0.30, 0.05) | 0.17 |
| BMI (kg/m<sup>2</sup>) | **13.4** | **13.6** | **0.11** | (-0.11, 0.33) | 0.32 | **13.1** | **12.9** | **-0.33** | (-0.62, -0.03) | 0.03 | **-0.04** | (-0.22, 0.13) | 0.65 |
| Waist circumference (cm) | **50.1** | **50.6** | **0.13** | (-0.09, 0.35) | 0.24 | **50.0** | **49.5** | **-0.38** | (-0.68, -0.08) | 0.01 | **-0.05** | (-0.22, 0.13) | 0.60 |
| Fat percentage (%) | **17.2** | **17.8** | **0.14** | (-0.08, 0.35) | 0.22 | **20.4** | **20.6** | **-0.01** | (-0.31, 0.29) | 0.93 | **0.09** | (-0.09, 0.26) | 0.34 |
| Fasting glucose (mmol/l) | **4.94** | **5.16** | **0.44** | (0.23, 0.65) | <0.001 | **4.77** | **4.93** | **0.31** | (0.01, 0.61) | 0.04 | **0.40** | (0.22, 0.57) | <0.001 |
| 30-min glucose (mmol/l) | **8.21** | **8.10** | **0.07** | (-0.16, 0.27) | 0.52 | **8.10** | **8.55** | **0.30** | (-0.03, 0.57) | 0.05 | **0.15** | (-0.05, 0.29) | 0.10 |
| 120-min glucose (mmol/l) | **5.27** | **5.49** | **0.24** | (0.03, 0.46) | 0.03 | **5.57** | **5.38** | **0.25** | (-0.05, 0.55) | 0.10 | **0.25** | (0.07, 0.42) | 0.006 |
| Fasting insulin (pmol/l) | **16.80** | **20.94** | **0.28** | (0.07, 0.49) | 0.01 | **20.04** | **18.84** | **-0.16** | (-0.46, 0.14) | 0.29 | **0.13** | (-0.05, 0.30) | 0.15 |
| 30-min insulin (pmol/l) | **138.0** | **131.7** | **0.06** | (-0.19, 0.23) | 0.60 | **163.3** | **153.7** | **0.06** | (-0.19, 0.41) | 0.70 | **0.06** | (-0.12, 0.22) | 0.52 |
| 120-min insulin (pmol/l) | **47.8** | **58.4** | **0.20** | (-0.05, 0.39) | 0.07 | **66.1** | **61.6** | **0.06** | (-0.27, 0.33) | 0.70 | **0.15** | (-0.06, 0.28) | 0.09 |
| HOMA-S | **262** | **218** | **-0.30** | (-0.51, -0.08) | 0.008 | **225** | **239** | **0.13** | (-0.17, 0.43) | 0.40 | **-0.15** | (-0.32, 0.03) | 0.10 |
| HOMA-β | **48** | **52** | **0.06** | (-0.16, 0.27) | 0.62 | **61** | **48** | **-0.23** | (-0.53, 0.06) | 0.12 | **-0.04** | (-0.22, 0.13) | 0.62 |
| Disposition Index (HOMA) | **143** | **119** | **-0.48** | (-0.51, 0.08) | <0.001 | **139** | **140** | **-0.11** | (-0.46, 0.15) | 0.48 | **-0.35** | (-0.47, -0.12) | <0.001 |
| Insulinogenic Index<sup>a</sup> | **1.57** | **1.54** | **-0.18** | (-0.40, 0.04) | 0.11 | **1.54** | **1.58** | **0.18** | (-0.12, 0.48) | 0.25 | **-0.06** | (-0.23, 0.12) | 0.53 |
| Matsuda Index<sup>b</sup> | **58.5** | **52.5** | **-0.27** | (-0.48, -0.05) | 0.01 | **46.8** | **51.5** | **0.03** | (-0.27, 0.33) | 0.83 | **-0.16** | (-0.33, 0.01)) | 0.07 |
| Disposition Index<sup>c</sup> | **6.8** | **6.7** | **-0.29** | (-0.51, 0.07) | 0.008 | **6.8** | **6.7** | **-0.04** | (-0.35, 0.29) | 0.77 | **-0.20** | (-0.37, -0.03) | 0.02 |
| **2 YEARS** |  |  |  |  |  |  |  |  |  |  |  |  |  |
| Height (cm) | **82.2** | **82.0** | **-0.13** | (-0.35, 0.09) | 0.24 | **81.3** | **79.6** | **-0.36** | (-0.66, -0.07) | 0.02 | **-0.21** | (-0.38, -0.04) | 0.02 |
| Weight (kg) | **9.9** | **9.8** | **-0.06** | (-0.28, 0.15) | 0.58 | **9.2** | **8.8** | **-0.45** | (-0.75, -0.16) | 0.003 | **-0.19** | (-0.37, -0.02) | 0.03 |
| **BIRTH** |  |  |  |  |  |  |  |  |  |  |  |  |  |
| Weight (g) | **2700** | **2700** | **-0.01** | (-0.23, 0.22) | 0.99 | **2550** | **2500** | **-0.19** | (-0.50, 0.11) | 0.20 | **-0.07** | (-0.25, 0.11) | 0.45 |
| Length (cm) | **48.2** | **47.8** | **-0.17** | (-0.39, 0.04) | 0.11 | **47.4** | **47.0** | **-0.41** | (-0.70, -0.11) | 0.007 | **-0.26** | (-0.43, -0.08) | 0.004 |
| Head circumference (cm) | **33.4** | **33.2** | **-0.17** | (-0.39, 0.05) | 0.12 | **32.7** | **32.5** | **-0.10** | (-0.40, 0.20) | 0.50 | **-0.14** | (-0.32, -0.03) | 0.10 |
| Gestation (days) | **273** | **272** | **-0.05** | (-0.27, 0.16) | 0.64 | **273** | **273** | **-0.19** | (-0.49, 0.1) | 0.20 | **-0.10** | (-0.27, 0.07) | 0.26 |
| **MOTHER** | **N=221** | **N=130** |  |  |  | **N=213** | **N=54** |  |  |  |  |  |  |
| Fasting glucose (mmol/l) in pregnancy (28-wk) | **3.9** | **4.0** | **0.14** | (-0.09, 0.38) | 0.23 | **3.8** | **3.8** | **0.07** | (-0.26, 0.40) | 0.68 | **0.11** | (-0.07, 0.30) | 0.23 |
|  |  |  | **Odds Ratio** | **95% CI** | **p** |  |  | **Odds Ratio** | **95% CI** | **p** | **Odds Ratio** | **95% CI** | **p** |
| Pregnancy glucose intolerance (FPG ≥ 5.1 mmol/l) | **14**<br>**(6.3%)** | **15**<br>**(11.5%)** | **1.91** | (0.89, 4.10) | 0.10 | **19**<br>**(8.9%)** | **9**<br>**(16.7%)** | **2.04** | (0.87, 4.81) | 0.11 | **1.97** | (1.11, 3.48) | 0.02 |
| Post-natal glucose intolerance (DM+IFG+IGT) | **62**<br>**(28.1%)** | **52**<br>**(39.7%)** | **1.67** | (1.01, 2.64) | 0.03 | **53**<br>**(24.9%)** | **17**<br>**(31.5%)** | **1.38** | (0.66, 2.61) | 0.33 | **1.58** | (1.08, 2.29) | 0.02 |
| Ever Overweight/Obese | **58**<br>**(26.2%)** | **23**<br>**(17.6%)** | **0.60** | (0.35, 1.02) | 0.06 | **46**<br>**(21.6%)** | **8**<br>**(14.8%)** | **0.63** | (0.28, 1.43) | 0.27 | **0.60** | (0.39, 0.96) | 0.03 |
| **FATHER** | **N=221** | **N=131** |  |  |  | **N=212** | **N=54** |  |  |  |  |  |  |
| Ever glucose intolerant (DM+IFG+IGT) | **113**<br>**(52.3%)** | **64**<br>**(48.9%)** | **0.91** | (0.59, 1.41) | 0.68 | **77**<br>**(36.2%)** | **30**<br>**(55.6%)** | **2.21** | (1.20, 4.04) | 0.01 | **1.23** | (0.87, 1.75) | 0.24 |
| Ever Overweight/Obese | **81**<br>**(36.7%)** | **44**<br>**(33.6%)** | **0.87** | (0.59, 1.54) | 0.56 | **71**<br>**(33.3%**<br>**)** | **19**<br>**(35.2%)** | **1.08** | (0.51, 1.90) | 0.78 | **0.94** | (0.65,<br>1.36) | 0.75 |

<sup>a</sup> $\ln\{\text{Insulin(30-minute/fasting)}/\text{Glucose(30-minute/fasting)}\}$ ;
<sup>b</sup> $10000/\sqrt{\{\text{Glucose fasting} \times \text{Insulin fasting} \times \text{mean glucose (F, 30min, 120min)} \times \text{mean insulin (F, 30min, 120min)}\}}$ (glucose in mmol/l; insulin in pmol/l);
<sup>c</sup> $\text{Insulinogenic index} + \ln(\text{Matsuda index})$ ;
<sup>d</sup> $(\text{HOMA-S} \times \text{HOMA-}\beta)/100$ ;
<sup>e</sup> $\text{HbA1C (mmol/mol)} = (10.93 \times \text{HbA1c(\%)}) - 23.5$ , source: NGSP's HbA1c converter at http://www.ngsp.org/convert1.asp;
<sup>s</sup> P-value calculated using chi-square test.
BMI: Body Mass Index; HOMA: Homeostatic Model Assessment models, SD: Standard deviation, SES: Socio economic status, SLI: Standard of Living Index.

### Ethics

The study was approved by village leaders and the KEM Hospital Research Centre Ethics Committee. Parents gave written consent; children under 18 years of age gave written assent, and written consent after reaching 18 years.

### Results

The analysis included 619 men and women with complete data (86% of the original live births). Mean BMI was 19.7 kg/m^2^ in men and 18.7 kg/m^2^ in women; 41% of men and 57% of women were underweight, and ∼10% were stunted (Supplemental Table S1). Eight percent of men and 4% of women were overweight/obese while 6% of men and 5% of women were centrally obese. Sixteen percent each of men and women were adipose (DXA). A total of 185 (30%) were glucose intolerant: one woman had diabetes, 37% of men and 20% of women had prediabetes. Men had more IFG (27%) than women (9%) but rates of IGT were similar (11% in both sexes). Thirty one percent of glucose intolerant men and 67% of glucose intolerant women were underweight. Glucose intolerant men, but not women, had higher BMI, fat% and waist circumference than NGT participants.

### Lifecourse evolution of glucose-insulin indices and comparison of glucose intolerant and NGT participants (Table 1 and Fig. 2)

Glucose intolerant participants had higher FPG than NGT participants at age 6, 12 (and 18) years and higher HbA1c at 15 years. Fasting insulin concentrations were similar at 6 and 12 years, but higher at 18 years in the glucose intolerant group. In NGT participants, insulin sensitivity indices (HOMA-S and Matsuda index) were the highest and insulin secretion indices (HOMA-B and insulinogenic index) lowest at 6 years of age; there was a progressive fall in insulin sensitivity and increase in insulin secretion from 6 to 18 years. In the glucose intolerant group, insulin sensitivity was highest and HOMA-B lowest at 6-years, and there was a fall in insulin sensitivity from 6- to 18-years accompanied by an increase in HOMA-B but not the insulinogenic index. The disposition index, however, showed a progressive fall from 6 to 18 years in both the NGT and the glucose intolerant groups, and was consistently lower in the glucose intolerant. Glucose intolerant men, but not women, had lower HOMA-S and Matsuda index at age 6 years.

Glucose intolerant men and women were shorter at birth, but there were no significant differences in birth weight compared to the NGT group. They continued to be shorter and lighter at 2 years, and women but not men, continued to be shorter and thinner until 6 years (Supplemental Fig. S2). Glucose intolerant men, but not women, gained more weight and BMI during adolescence than the NGT group.

### Parental influences

Glucose intolerant men and women were more likely than the NGT group to have a mother with glucose intolerance in pregnancy or post-natally, and a mother who was not overweight or obese. Glucose intolerant women were also more likely to have a father with glucose intolerance. There was no difference in the duration of exclusive or total breastfeeding in the two groups.

### Multivariate modelling of glucose intolerance at 18 years (Table 2)

Significant predictors in both sexes were maternal pregnancy glucose intolerance, a mother who had never been overweight/obese, and 6-year and 12-year FPG. Women had a lower incidence, and an additional association with paternal glucose intolerance. Apart from smaller length or head circumference at birth, none of the childhood growth variables were significantly related (examined using conditional BMI and height gain through childhood; data not shown in Table 2). Greater adiposity at 18 years was associated with an increased risk only among men. SES was not related to 18-year glucose intolerance.

**Table 2:**
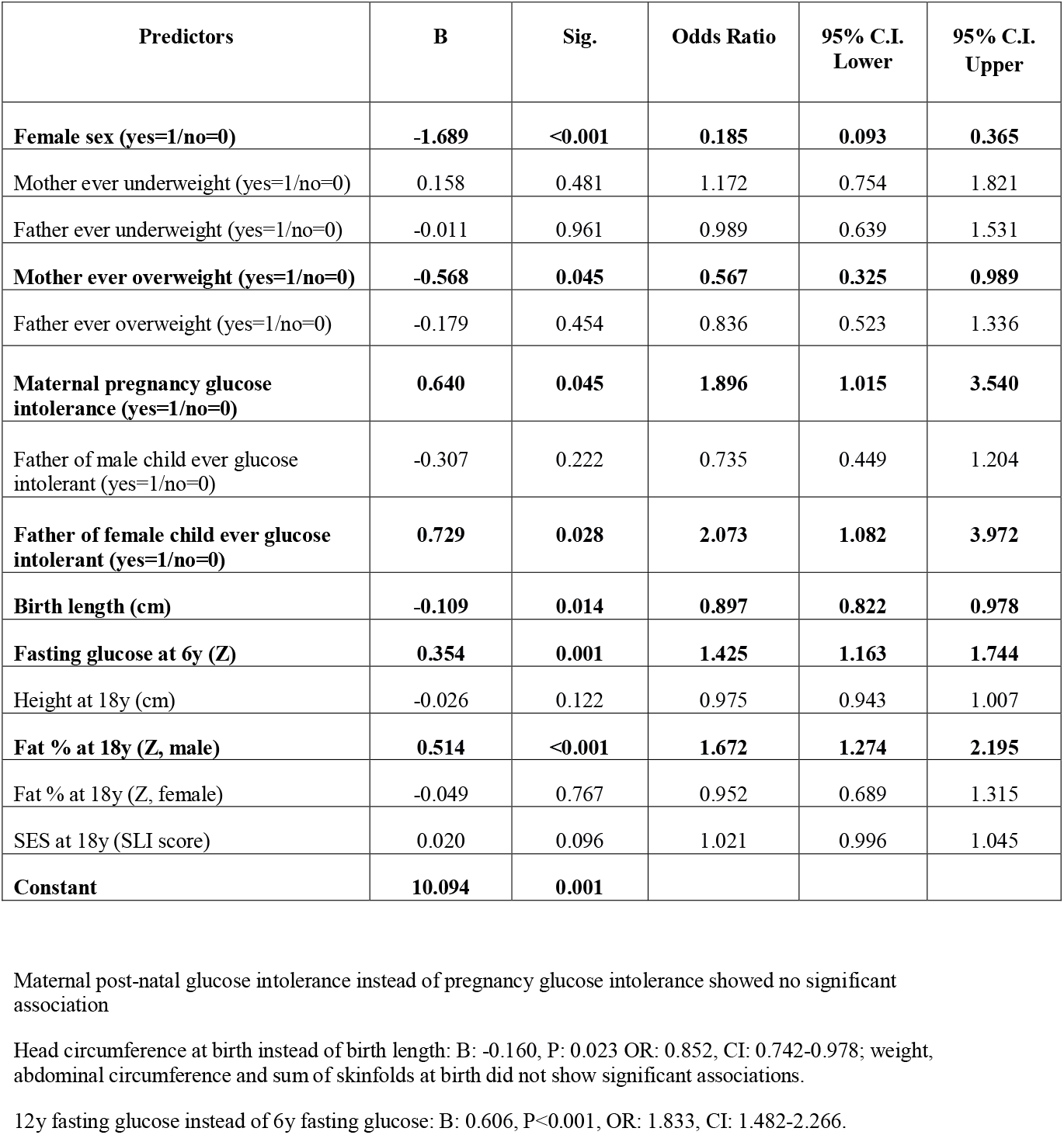
Multivariate regression with the outcome of glucose intolerance at age 18 years.

We examined these associations separately in IFG and IGT groups (Supplemental Tables S2a and S2b) aware that this analysis has limited power. Both groups were small at birth and had a reduced beta cell compensatory response in childhood compared to NGT participants.

### Childhood and adolescent FPG as predictors of later glucose intolerance

We further investigated the associations between FPG at 6 and 12 years and glucose intolerance at 18 years. The prevalence was 2.5 times higher among those in the highest quintile of FPG at 6 years, and 4.0 times higher at 12 years, than among those in the lowest quintile (Fig. 3). ROC curves (Supplemental Figs. S3a and S3b) for 18-year glucose intolerance showed that the area under the curve (AUC) using FPG was 0.658 at age 6-years and 0.700 at age 12 years (adjusted for sex). These values increased marginally to 0.686 and 0.723 respectively when the model included all the predictors in Table 2.

**Figure 2:**
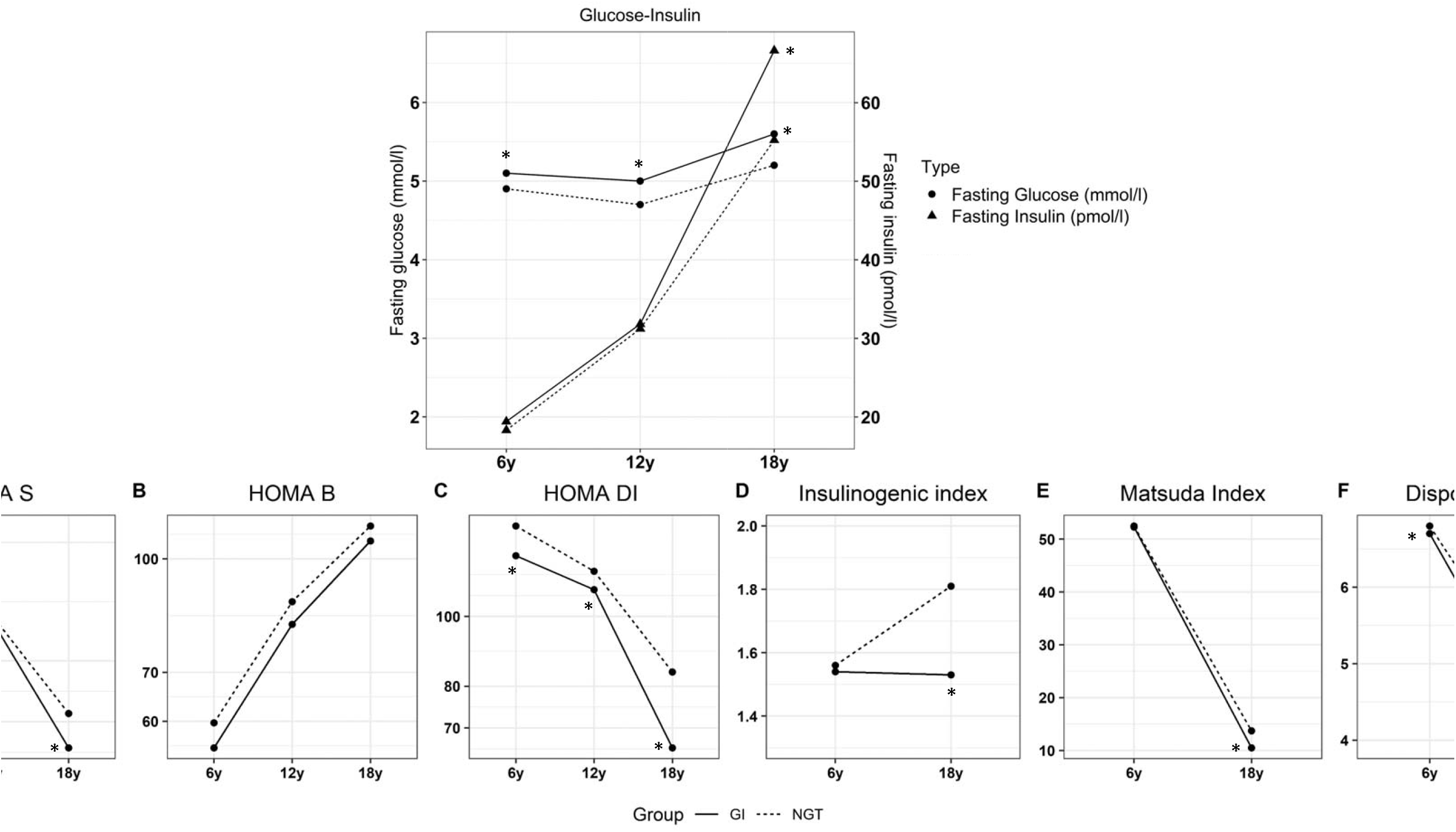
Lifecourse evolution of glucose-insulin metabolism in participants of PMNS [NGT vs Glucose intolerant]. The figure shows the lifecourse evolution of parameters of glucose and insulin metabolism in NGT (dotted line) and glucose intolerant (solid line) participants. The top panel shows fasting plasma glucose (mmol/l) and fasting plasma insulin (pmol/l). The bottom panel shows HOMA indices (A-C) and dynamic indices (D-F). Significant differences between the two groups (p<0.05) are indicated by an asterisk.

**Figure 3:**
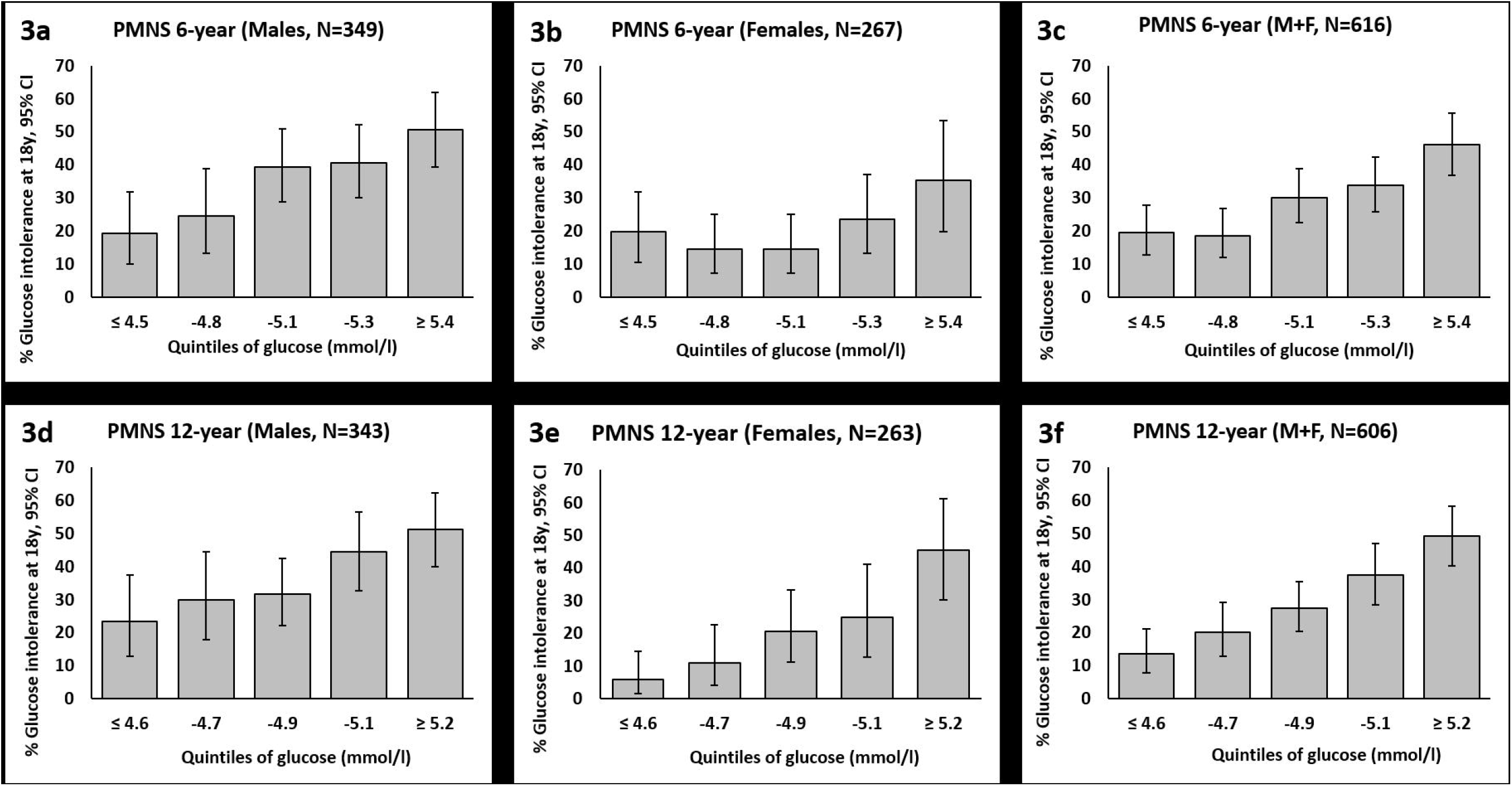
Probability of glucose intolerance at 18 years according to childhood fasting glucose. The prevalence of glucose intolerance at 18 years according to quintiles of fasting plasma glucose concentration at 6-years (3a, 3b, 3c) and 12-years (3d, 3e, 3f).

We replicated this analysis in the two other cohorts. In the PCS (N=355), 66 (19%) participants were glucose intolerant at age 21 years (5 type 2 diabetes + 40 IFG + 21 IGT). They had higher BMI, and lower insulin sensitivity and insulin secretion than the NGT participants. They also had higher FPG concentrations (4.8 v 4.6 mmol/l, p=0.026) and lower disposition index at 8 years (12.9 v 15.3, p=0.026). The prevalence of glucose intolerance at 21 years was 1.7 times higher for those in the highest tertile of 8-year FPG than among those in the lowest tertile (Supplemental Fig. S4a). In the Extended PMNS cohort, FPG at age 12 years, but not 6 years, was positively associated with higher FPG concentration at 18 years. The likelihood of being in highest tertile of FPG at 18 years was 3.1 times higher in those in the highest tertile of FPG at 12 years than among those in the lowest tertile (Supplemental figure S4b).

## Discussion

We found a high prevalence of glucose intolerance in this young thin rural Indian cohort, higher in men than women. The glucose intolerant participants had higher glucose from early childhood compared to NGT, reflecting an inadequate compensatory insulin response to decreasing insulin sensitivity. Our novel intergenerational lifecourse analysis revealed other factors associated with adult glucose intolerance, including parental glucose intolerance and reduced fetal and infant growth. These findings support an intergenerational DOHaD model of type 2 diabetes, which was first conceptualised in the ‘thrifty phenotype’ hypothesis, which attributed adult glucose intolerance to a fetus having to be metabolically thrifty in order to survive intra-uterine nutritional deprivation. (9) These ideas challenge the predominant paradigm that type 2 diabetes is a disorder of β-cell decompensation due to adult obesity-related insulin resistance. (23)

### Childhood glucose predicts adult glucose intolerance

Higher childhood and adolescent FPG concentrations were strong and graded predictors of future glucose intolerance, which was predicted with 66% and 70% confidence by this measure alone at 6 and 12 years respectively. These findings provide a simple biomarker for future risk. Our results demonstrate, for the first time in humans, continuous tracking of glycemia from childhood to adulthood and a strong predictive value of childhood FPG for later glucose intolerance. We were able to validate the prediction in two other cohorts in Pune. The Bogalusa and i3C studies hinted at tracking from a single childhood timepoint and the Early Bird study showed tracking from early childhood into adolescence. (24-26) These results should convince paediatricians to measure glucose concentrations in children, and policy makers to promote preventive measures at a younger age.

### Parental factors

Intergenerational influences on glucose intolerance appear to reflect a combination of factors. Genetic factors are obviously important, but a specific influence of pregnancy glycemia suggests an epigenetic programming effect. It is intriguing that a lack of overweight-obesity in the mother was associated with glucose intolerance in the child. The parents and grand-parents of our cohort grew up in an impoverished drought-prone area. The mothers were short (mean height 1.52 m), thin (mean BMI 18.1 kg/m^2^) and had low macro- and micro-nutrient intakes and heavy physical workloads in pregnancy. (13) Maternal glucose concentrations in pregnancy were relatively low and few had GDM, but were nevertheless associated with glucose intolerance in their offspring. Our results suggest that the current epidemic of diabetes in young Indians may be rooted in ‘dual teratogenesis’ i.e., simultaneous intrauterine exposure to multiple nutritional deficits and (minimally) elevated maternal glucose. (27) Differences in duration of breast feeding do not seem to have played a role. Sex-specific effects of paternal glycaemia suggest a role for imprinting, and merit further investigation. (28)

### Growth and sex

Rather than low birth weight, short birth length and small head circumference were associated with adult glucose intolerance. While an association of short birth length with later diabetes has been described in another Indian cohort, (29) that with smaller head circumference is a new finding. Human intra-uterine growth is governed by the necessity to maintain brain growth (‘brain sparing’), and our finding of smaller head circumference in the glucose intolerant suggests a relatively severe nutritional challenge. Circulatory adjustments for brain sparing are likely to compromise the development of important abdominal organs. (30) Of relevance to glucose intolerance, detrimental effects of intra-uterine under-nutrition on the structure and function of the liver and pancreas have been well demonstrated in animal models. (31,32)

Glucose intolerant men and women showed different post-natal growth trajectories (Supplemental figure S2). Women remained shorter and thinner, and two-thirds of glucose intolerant women were underweight at 18 years. Glucose intolerant men gained more body mass during puberty than the NGT group. There were similar findings in the Delhi and Helsinki birth cohorts, which showed that small size in infancy but greater childhood and adolescent weight gain were associated with later glucose intolerance. (10,11). It is noteworthy that a third of the glucose intolerant men were still underweight (low BMI) though more adipose (body fat%) than the NGT group. These findings support our previous observations of the ‘thin-fat’ Indian phenotype predisposing to diabetes. (30) Becoming heavy ‘relative to oneself’ (upwards centile crossing) is a strong risk factor for diabetes in those who were growth restricted in early life. (10,11,21).

We propose that type 2 diabetes in Indians has its roots in a history of multi-generational undernutrition, leaving a legacy of fetal growth restriction, combined with recent rapid nutritional transition which places increased metabolic demands on developmentally stunted metabolic systems. Between 1830 and 1980, Indians failed to gain height while Europeans gained up to 15 cm. (33) The reasons for the dramatic historical failure of height gain in Indians can only be environmental stresses: famines, undernutrition, and infectious disease. Children in our study, in contrast, are on average five cm taller and five kg heavier than their parents, suggesting a recent rapid transition. The drivers of such changes in our study area include a reliable water supply from a dam (supporting irrigation and cash-crop farming), a new industrial estate (generating paid employment), and improved literacy rates. The sex difference in glucose intolerance may also partly be due to societal preferences for the male child.

### Pathophysiology

A typical type 2 diabetes patient demonstrates both reduced insulin secretion and sensitivity, with varying contributions in different patients. There are only a few lifecourse studies, predominantly from the ‘obese’ western world which showed that higher childhood FPG, BMI, insulin concentrations and HOMA-IR were predictors of future glucose intolerance; it is noteworthy that HOMA-β and DI were not mentioned. (23, 24) The role of reduced β-cell secretion relative to insulin insensitivity (DI) was stressed in the Early Bird study and the ADA statement on youth onset type 2 diabetes. (34,35) Our data highlights that insulin sensitivity progressively decreases from childhood into adulthood, accompanied by an increase in beta cell secretion in the NGT group evident in both fasting and stimulated states, indicating good beta cell reserve. In contrast, in the glucose intolerant there was little increase in stimulated insulin secretion. A progressive decrease in the disposition index captured the inadequacy of compensatory beta cell secretion in the face of decreasing insulin sensitivity. Consistently lower disposition index in the glucose intolerant group suggests an underperforming beta cell from early life. Our results highlight the importance of interpreting insulin secretion in relation to prevailing insulin sensitivity to detect relevant pathophysiology. We believe this is a novel description of the evolution of glucose-insulin physiology during childhood, puberty and young adulthood.

Most previous studies, including some of ours, have considered insulin insensitivity the primary driver of diabetes, probably because of inadequate investigation of insulin secretion. The importance of diminished insulin secretion in the pathophysiology of type 2 diabetes in Indians has recently been highlighted (36). Severely insulin deficient diabetes (SIDD) was the most common sub-type in our young (<45 years) type 2 diabetic patients (submitted for publication), and also in the migrant Indians in the USA (37). In contrast, in a Swedish cohort, the main subtype was Mild Obese Diabetes (MOD).

### Implications

The strong prediction of adult glucose intolerance from childhood glucose measurements mandates the monitoring of children’s plasma glucose concentrations. Our research will help identify at-risk individuals in childhood and potentially reduce risk using therapies which improve insulin secretion and sensitivity. Measurement of birth length and head circumference in addition to weight would add to risk prediction. Persistently higher glucose levels from early childhood, even within the normal range, have the potential to epigenetically affect ova and sperm, contributing to a higher risk of diabetes in the next generation. Thus, early identification and management of at-risk individuals could benefit future generations. Our findings may be relevant to other developing populations with a history of nutritional deprivation.

Strengths of our study are exceptional follow up over 20 years (92% of survivors), longitudinal anthropometry from birth, and serial glucose-insulin data from childhood. All measurements used uniform methods throughout, and serial insulin assays were calibrated against the same international reference. Participants included were comparable to those excluded at each stage (Supplemental figure S1). The PMNS findings were validated in a rural as well as an urban cohort, increasing their generalisability, though we used a more arbitrary cut-point (highest tertile) for glucose intolerance in one validation cohort due to small numbers with prediabetes. Limitations were that for logistic reasons, we used ‘epidemiological’ rather than ‘gold standard’ measures of insulin action and secretion, though the former are well accepted and used widely in cohort studies; and at 12 years we had only fasting glucose-insulin measurements.

In well-nourished Europeans, experimental starvation causes acute glucose intolerance (39). We took care to avoid any ‘starvation’ among our participants in the week before the OGTT. In addition, elevated FPG and HbA1c many years earlier suggest ongoing long-term hyperglycemia. The predominance of thin SIDD patients in our urban diabetes clinics further supports chronic undernutrition as an underlying etiological factor. Therefore, type 2 diabetes in undernourished and transitioning populations may be the new avatar of malnutrition-related diabetes (MRDM), a previously recognised sub-class of diabetes that fell into obscurity due to a lack of prospective data and an increasing focus on obesity-related diabetes. (40)

In conclusion, glucose intolerance in thin young rural Indian adults is heralded by slower skeletal and brain growth *in utero*, and impaired compensatory insulin secretion and higher glycaemia from childhood. In men, pubertal weight gain aggravated insulin insensitivity and glucose intolerance. Glucose intolerance was seen in women despite continued undernutrition. We describe novel interactions between beta cell secretory capacity and age-related insulin insensitivity in an undernourished population leading to glucose intolerance at young age. Our findings reveal the pitfalls of cross-sectional studies in adults to postulate antecedent events, and stress the importance of prospective life-course measurements.

## Data Availability

Data is available with Prof C S Yajnik for sharing to confirm our findings and for additional analyses by applying to the corresponding author with a 200 word plan of analysis. Data sharing is subject to KEMHRC Ethics Committee approval and Government of India Health Ministry advisory committee permission.

## Acknowledgements

We are grateful to all study participants and their family members for co-operation over many years. We thank the late Prof DJP Barker, Dr B Coyaji, and Dr VN Rao for their support in establishing the PMNS. We thank Dr VS Padbidri and Dr L Garda, former and current Directors of Research, KEM Hospital Research Centre. We also thank the staff of the Diabetes Unit for their help in conducting the study, particularly Drs S Hirve, N Joshi, U Deshmukh, A Bavdekar, H Lubree, and R Ladkat, N Memane, C Joglekar, S Bagate, A Bhalerao, S Chaugule, R Dendge, T Deokar, M Gaikwad, N Gurav, S Jagtap, J Kalokhe, S Pandit, F Rajgara, D Raut, L Ramdas, M Raut, R Saswade, and V Solat. We thank Dr SS Naik, head of biochemistry, KEM Hospital for assay standardization. We are grateful to the Indian Council of Medical Research, the Department of Biotechnology, India, the Wellcome Trust and Medical Research Council, UK for their funding support. We acknowledge the support of Dr N D Deshmukh and the Zilla Parishad, Pune. Prof CS Yajnik is guarantor of this work, takes responsibility of data integrity, accuracy and had access to all data.

## Authors’ contributions

CY and CF conceptualized the study. SB, AB, RW and CO were involved in statistical analysis. KJC, PY, KK, AP, SB contributed to conduct of the study. CY, SP, CF, CO prepared the manuscript. DB was involved in laboratory measurements.

## Data sharing statement

Data is available with Prof C S Yajnik for sharing to confirm our findings and for additional analyses by applying to the corresponding author with a 200-word plan of analysis. Data sharing is subject to KEMHRC Ethics Committee approval and Government of India’s Health Ministry advisory committee permission.

## References

1. India State-Level Disease Burden Initiative Diabetes Collaborators. The increasing burden of diabetes and variations among the states of India: the Global Burden of Disease Study 1990- 2016. Lancet Glob Health. 2018; 6:e1352–e1362.

2. Comprehensive National Nutrition Survery (2016- 2018) reports. Ministry of Health and Family Welfare, Government of India, Population Council and UNICEF. (https://nhm.gov.in/index1.php?lang=1&level=2&sublinkid=1332&lid=713, last accessed September 2020).

3. Yajnik CS. The insulin resistance epidemic in India: fetal origins, later lifestyle, or both? Nutr Rev. 2001; 59:1–9.

4. Yajnik CS, Shelgikar KM, Naik SS, et al. Impairment of glucose tolerance over 10 years in middle-aged normal glucose tolerant Indians. Diabetes Care. 2003; 26:2212–3.

5. Ramachandran A, Ma RC, Snehalatha C. Diabetes in Asia. Lancet. 2010 Jan 30; 375:408–18.

6. Anjana RM, Deepa M, Pradeepa R, et al. Prevalence of diabetes and prediabetes in 15 states of India: results from the ICMR-INDIAB population-based cross-sectional study. Lancet Diabetes Endocrinol. 2017; 5:585–596.

7. Hales CN, Barker DJ, Clark PM, et al. Fetal and infant growth and impaired glucose tolerance at age 64. BMJ. 1991; 303:1019–1022.

8. Whincup PH, Kaye SJ, Owen CG, et al. Birthweight and risk of type 2 diabetes: a quantitative systematic review of published evidence. JAMA. 2008; 300: 2885–97.

9. Hales CN, Barker DJ. Type 2 (non-insulin-dependent) diabetes mellitus: the thrifty phenotype hypothesis. Diabetologia. 1992; 35:595–601.

10. Bhargava SK, Sachdev HS, Fall CH, et al. Relation of serial changes in childhood body-mass index to impaired glucose tolerance in young adulthood. N Engl J Med. 2004; 350:865–75.

11. Eriksson JG, Osmond C, Kajantie E, Forsén TJ, Barker DJ. Patterns of growth among children who later develop type 2 diabetes or its risk factors. Diabetologia. 2006; 49:2853–8.

12. Tuomilehto J, Lindström J, Eriksson JG, et al. Prevention of type 2 diabetes mellitus by changes in lifestyle among subjects with impaired glucose tolerance. N Engl J Med. 2001; 3:1343–50.

13. Rao S, Yajnik CS, Kanade A, et al. Intake of micronutrient-rich foods in rural Indian mothers is associated with the size of their babies at birth: Pune Maternal Nutrition Study. J Nutr. 2001; 131:1217–24.

14. Joglekar CV, Fall CH, Deshpande VU, et al. Newborn size, infant and childhood growth, and body composition and cardiovascular disease risk factors at the age of 6 years: the Pune Maternal Nutrition Study. Int J Obes (Lond). 2007; 31:1534–44.

15. Matsuda M, DeFronzo RA. Insulin sensitivity indices obtained from oral glucose tolerance testing: comparison with the euglycemic insulin clamp. Diabetes Care. 1999; 22:1462–70.

16. DeFronzo RA, Matsuda M. Reduced time points to calculate the composite index. Diabetes Care. 2010; 33:e93.

17. Wareham NJ, Phillips DI, Byrne CD, at al. The 30 minute insulin incremental response in an oral glucose tolerance test as a measure of insulin secretion. Diabet Med. 1995 ;12:931.

18. Bergman RN, Ader M, Huecking K, Van Citters G. Accurate assessment of beta-cell function: the hyperbolic correction. Diabetes. 2002 ;51:S212–20.

19. National Family Health Survey (NFHS-4), 2015-16: India. International Institute for Population Sciences (IIPS) and ICF. 2017. (http://rchiips.org/nfhs/NFHS-4Reports/India.pdflast, accessed September 2020)

20. American Diabetes Association. 2. Classification and Diagnosis of Diabetes: Standards of Medical Care in Diabetes-2019. Diabetes Care. 2019; 42:S13–S28.

21. Metzger BE, Gabbe SG, Persson B et al. International association of diabetes and pregnancy study groups recommendations on the diagnosis and classification of hyperglycemia in pregnancy. Diabtes Care. 2010; 33:676–682.

22. Bavdekar A, Yajnik CS, Fall CH, Bapat S, Pandit AN, Deshpande V, Bhave S, Kellingray SD, Joglekar C. Insulin resistance syndrome in 8-year-old Indian children: small at birth, big at 8 years, or both? Diabetes. 1999; 48:2422–9.

23. Kahn SE, Cooper ME, Del Prato S. Pathophysiology and treatment of type 2 diabetes: perspectives on the past, present, and future. Lancet. 2014; 383:1068–83.

24. Nguyen QM, Srinivasan SR, Xu JH, et al. Utility of childhood glucose homeostasis variables in predicting adult diabetes and related cardiometabolic risk factors: the Bogalusa Heart Study. Diabetes Care. 2010; 33:670–5.

25. Hu T, Jacobs DR Jr, Sinaiko AR, Bazzano LA, Burns TL, Daniels SR, et al. Childhood BMI and Fasting Glucose and Insulin Predict Adult Type 2 Diabetes: The International Childhood Cardiovascular Cohort (i3C) Consortium. Diabetes Care. 2020; 43:2821–2829.

26. Jeffery AN, Metcalf BS, Hosking J, Streeter AJ, Voss LD, Wilkin TJ. Age before stage: insulin resistance rises before the onset of puberty: a 9-year longitudinal study (EarlyBird 26). Diabetes Care. 2012; 35:536–41.

27. Yajnik CS. Nutrient-mediated teratogenesis and fuel-mediated teratogenesis: two pathways of intrauterine programming of diabetes. Int J Gynaecol Obstet. 2009; 104 Suppl 1:S27–31.

28. Su L, Patti ME. ME Nongenetic Intergenerational Transmission of Metabolic Disease Risk. Curr Diab Rep. 2019; 19:38.

29. Fall CH, Stein CE, Kumaran K, Cox V, Osmond C, Barker DJ, Hales CN. Size at birth, maternal weight, and type 2 diabetes in South India. Diabet Med. 1998; 15:220–7.

30. Yajnik CS. CS epidemic in India: intrauterine origins? Proc Nutr Soc. 2004; 63:387–96.

31. Hoet JJ, Ozanne S, Reusens B. Influences of pre- and postnatal nutritional exposures on vascular/endocrine systems in animals. Environ Health Perspect. 2000; 108 Suppl 3:563–8.

32. Hardikar AA, Satoor SN, Karandikar MS, et al. Multigenerational Undernutrition Increases Susceptibility to Obesity and Diabetes that Is Not Reversed after Dietary Recuperation. Cell Metab. 2015; 22:312–9

33. NCD Risk Factor Collaboration (NCD-RisC). A century of trends in adult human height. Elife. 2016; 5:e13410.

34. Arslanian S, Bacha F, Grey M, Marcus MD, White NH, Zeitler P. Evaluation and Management of Youth-Onset Type 2 Diabetes: A Position Statement by the American Diabetes Association. Diabetes Care. 2018; 41:2648–2668.

35. Hosking J, Metcalf BS, Jeffery AN, Streeter AJ, Voss LD, Wilkin TJ. TJ of early beta-cell deficiency among children who show impaired fasting glucose: 10-yr cohort study (EarlyBird 56). Pediatr Diabetes. 2013; 14:481–9.

36. Mohan V, Amutha A, Ranjani H, Unnikrishnan R, Datta M, Anjana RM, Staimez L, Ali MK, Narayan KM. KM of β-cell function and insulin resistance with youth-onset type 2 diabetes and prediabetes among Asian Indians. Diabetes Technol Ther. 2013; 15:315–22.

37. Bancks MP, Bertoni AG, Carnethon M, et al. Association of Diabetes Subgroups with Race/Ethnicity, Risk Factor Burden and Complications: The MASALA and MESA Studies [published online ahead of print, 2021 Jan 27]. J Clin Endocrinol Metab. 2021; dgaa962. doi:10.1210/clinem/dgaa962

38. Fleming TP, Watkins AJ, Velazquez MA, et al. Origins of lifetime health around the time of conception: causes and consequences. Lancet. 2018; 391:1842–1852.

39. Göschke H. H of glucose intolerance during fasting: differences between lean and obese subjects. Metabolism. 1977; 26:1147–1153.

40. Abu-Bakare A, Taylor R, Gill GV, et al. Tropical or malnutrition-related diabetes: a real syndrome? Lancet. 1986; 1:1135–8.

