## Supplementary material for "Poor *in-utero* growth, and reduced beta cell compensation and high fasting glucose from childhood, are harbingers of glucose intolerance in young Indians": -

**Supplemental material: Index**

| Sr No. | Item | Pg. No. |
|---|---|---|
| 1 | **Supplemental table S1:**<br><br>Cohort characteristics at 18, 12, 6 and 2 years, and newborn, infant and parent measurements | 3-7 |
| 2 | **Supplemental table S2a:**<br><br>Lifecourse data in participants with NGT, IFG, IGT and both IFG+IGT groups (Males) | 8-10 |
| 3 | **Supplemental table S2b:**<br><br>Lifecourse data in participants with NGT, IFG, IGT and both IFG+IGT groups (Females) | 11-13 |
| 4 | **Supplemental table S3:**<br><br>Associations of lifecourse measurements with IFG and IGT, each compared with NGT, at 18 years | 14-16 |
| 5 | **Supplemental table S4:**<br><br>Methods for anthropometry and body composition measurements in children | 17 |
| 6 | **Supplemental table S5:**<br><br>Quality assessments for glucose and insulin measurements in the Pune Maternal Nutrition Study (1993-2013) | 18-19 |
| 7 | **Supplemental figure S1:**<br><br>STROBE flow diagram for the Pune Maternal Nutrition Study. | 20-21 |
| 8 | **Supplemental figure S2:** | 22 |

| | | |
|---|---|---|
| | Comparative sizes across lifecourse of participants with glucose intolerance | |
| **9** | **Supplemental figure S3:**<br><br>Predictive probability of glucose intolerance at 18 years in PMNS | **23** |
| **10** | **Supplemental figure S4:**<br><br>Glucose intolerance in the PCS and Extended PMNS cohorts | **24** |

**Supplemental Table S1: Cohort characteristics at 18, 12, 6 and 2 years, and newborn, infant and parent measurements**

| Measure | Statistic | MEN (n=352) | | WOMEN (n=267) | |
|---|---|---|---|---|---|
| **18 YEARS** | | | | | |
| Age (years) | Mean (SD) | **18.2** | (0.5) | **17.7** | (0.6) |
| Height (cm) | Mean (SD) | **169.6** | (6.7) | **156.9** | (5.9) |
| Stunted (WHO z<-2) | N (%) | **33** | (9.4) | **28** | (10.5) |
| Weight (kg) | Mean (SD) | **56.8** | (10.7) | **46.0** | (7.8) |
| Body Mass Index (BMI, $kg/m^2$) | Mean (SD) | **19.7** | (3.3) | **18.7** | (3.1) |
| Underweight [a] (BMI<18.5 $kg/m^2$) | N (%) | **146** | (41.5) | **151** | (56.6) |
| Overweight/Obese [b] (BMI>25.0 $kg/m^2$) | N (%) | **27** | (7.7) | **11** | (4.1) |
| Overweight/Obese (BMI>23.0 $kg/m^2$) | N (%) | **53** | (15.1) | **19** | (7.1) |
| Waist circumference (cm) | Mean (SD) | **72.9** | (9.1) | **68.0** | (6.8) |
| Central obesity[c] | N (%) | **21** | (6.0) | **13** | (4.9) |
| Fat mass (kg) | Median (LQ, UQ) | **7.1** | (4.4, 11.8) | **12.1** | (9.3, 15.2) |
| Lean mass (kg) | Mean (SD) | **44.5** | (5.4) | **30.4** | (3.5) |
| Fat percentage (%) | Median (LQ, UQ) | **13.1** | (8.9, 20.4) | **28.0** | (22.9, 31.9) |

| | | | | | |
|---|---|---|---|---|---|
| Adipose[d] | N (%) | **56** | (16.1) | **41** | (15.5) |
| Fasting glucose (mmol/l) | Median (LQ, UQ) | **5.4** | (5.2, 5.6) | **5.2** | (5.0, 5.3) |
| 30-min glucose (mmol/l) | Median (LQ, UQ) | **8.2** | (7.5, 9.4) | **8.3** | (7.5, 9.0) |
| 120-min glucose (mmol/l) | Median (LQ, UQ) | **6.1** | (5.3, 7.0) | **6.3** | (5.5, 7.0) |
| Fasting insulin (pmol/l) | Median (LQ, UQ) | **52.8** | (39.0, 75.8) | **63.6** | (49.2, 81.0) |
| 30-min insulin (pmol/l) | Median (LQ, UQ) | **477.3** | (331.1, 707.9) | **579.0** | (391.2, 768.6) |
| 120-min insulin (pmol/l) | Median (LQ, UQ) | **280.2** | (177.0, 423.6) | **392.4** | (249.6, 579.0) |
| Impaired fasting glucose (IFG)[e] | N (%) | **94** | (26.7) | **23** | (8.6) |
| Impaired gluc tolerance (IGT)[f] | N (%) | **37** | (10.5) | **30** | (11.2) |
| Diabetes[g] | N (%) | **0** | (0) | **1** | (0.4) |
| Glucose Intolerant (DM+IFG+IGT) | N (%) | **131** | (37.2) | **54** | (20.2) |
| HOMA-S | Median (LQ, UQ) | **86** | (62, 118) | **73** | (58, 95) |
| HOMA-β | Median (LQ, UQ) | **94** | (75, 115) | **115** | (94,133) |
| Insulinogenic index[h] | Median (LQ, UQ) | **1.7** | (1.4, 2.1) | **1.7** | (1.4, 2.1) |
| Matsuda index[i] | Median (LQ, UQ) | **13.3** | (9.6, 19.2) | **11.6** | (8.7, 14.9) |
| Disposition index (HOMA)[j] | Median (LQ, UQ) | **79** | (68, 92) | **84** | (74, 95) |
| Disposition index[k] | Median (LQ, UQ) | **4.3** | (3.9, 4.9) | **4.1** | (3.7, 4.6) |
| **12 YEARS** | | | | | |

| | | | | | |
|---|---|---|---|---|---|
| Height (cm) | Mean (SD) | **138.7** | (8.5) | **140.2** | (8.4) |
| BMI ($kg/m^2$) | Mean (SD) | **14.9** | (1.7) | **14.7** | (2.2) |
| Waist (cm) | Mean (SD) | **58.1** | (5.4) | **56.3** | (5.3) |
| Fat percentage (%) | Median (LQ, UQ) | **13.8** | (11.1, 17.4) | **17.5** | (14.3, 22.2) |
| Fasting glucose (mmol/l) | Median (LQ, UQ) | **4.9** | (4.6, 5.1) | **4.7** | (4.5, 5.1) |
| Fasting insulin (pmol/l) | Median (LQ, UQ) | **28.2** | (18.0, 40.8) | **34.0** | (24.5, 47.2) |
| HOMA-S | Median (LQ, UQ) | **161** | (116, 249) | **138** | (101, 190) |
| HOMA-β | Median (LQ, UQ) | **72** | (51, 92) | **88** | (71, 108) |
| Disposition index (HOMA) | Median (LQ, UQ) | **120** | (102, 141) | **120** | (102, 140) |
| **6 YEARS** | | | | | |
| Height (cm) | Mean (SD) | **110.3** | (4.8) | **109.3** | (4.3) |
| BMI ($kg/m^2$) | Mean (SD) | **13.5** | (0.8) | **13.2** | (0.9) |
| Waist (cm) | Mean (SD) | **50.4** | (2.6) | **50.1** | (2.6) |
| Fat percentage (%) | Median (LQ, UQ) | **17.5** | (13.9, 20.5) | **20.4** | (17.6, 23.9) |
| Fasting glucose (mmol/l) | Median (LQ, UQ) | **5.1** | (4.7, 5.3) | **4.8** | (4.6, 5.2) |
| 30-min glucose (mmol/l) | Median (LQ, UQ) | **8.2** | (6.9, 9.3) | **8.1** | (7.2, 9.2) |
| 120-min glucose (mmol/l) | Median (LQ, UQ) | **5.4** | (4.6, 6.2) | **5.6** | (4.9, 6.3) |
| Fasting insulin (pmol/l) | Median (LQ, UQ) | **17.9** | (9.1, 27.3) | **19.6** | (9.7, 30.8) |
| 30-min insulin (pmol/l) | Median (LQ, UQ) | **136.1** | (86.1, 197.9) | **161.5** | (18,37) |

| | | | | | |
|---|---|---|---|---|---|
| 120-min insulin (pmol/l) | Median (LQ, UQ) | **49.9** | (27.2, 81.6) | **64.8** | (37.6, 103.7) |
| HOMA-S | Median (LQ, UQ) | **249** | (167, 448) | **228** | (149, 440) |
| HOMA-β | Median (LQ, UQ) | **50** | (33, 71) | **60** | (37, 81) |
| Insulinogenic index[h] | Median (LQ, UQ) | **1.6** | (1.1, 2.1) | **1.5** | (1.1, 2.1) |
| Matsuda index[i] | Median (LQ, UQ) | **55.4** | (36.0, 80.2) | **47.5** | (32.7, 69.2) |
| Disposition index (HOMA)[j] | Median (LQ, UQ) | **129** | (105, 157) | **137** | (107, 173) |
| Disposition index[k] | Median (LQ, UQ) | **6.8** | (6.4, 7.1) | **6.8** | (6.5, 7.1) |
| **2 YEARS** | | | | | |
| Height (cm) | Mean (SD) | **82.3** | (3.4) | **80.8** | (3.2) |
| Weight (kg) | Mean (SD) | **9.9** | (1.1) | **9.2** | (1.1) |
| **BIRTH** | | | | | |
| Birth weight (g) | Mean (SD) | **2700** | (365) | **2550** | (371) |
| Birth length (cm) | Mean (SD) | **47.8** | (2.4) | **47.1** | (2.2) |
| Head circumference (cm) | Mean (SD) | **33.3** | (32.3, 34.1) | **32.7** | (31.8, 33.5) |
| Gestation (days) | Mean (SD) | **271** | (12) | **272** | (11) |
| SGA | N (%) | **169** | (48) | **134** | (50.2) |
| **MOTHER (pre-pregnancy)** | | **N=352** | | **N=267** | |
| Height (cms) | Mean (SD) | **151.9** | (5.03) | **151.8** | (4.8) |
| BMI (kg/m$^2$) | Mean (SD) | **18.1** | (1.8) | **17.9** | (1.9) |

| **FATHER (during wife's pregnancy)** | | **N=334** | | **N=254** | |
|---|---|---|---|---|---|
| Height (cms) | Mean (SD) | **164.5** | (6.6) | **165.6** | (5.3) |
| BMI (kg/m$^2$) | Mean (SD) | **21.0** | (3.2) | **20.7** | (3.2) |

[a]BMI <18.5 kg/m$^2$; [b]BMI ≥25 kg/m$^2$; [c]Waist circumference ≥90 cm in men and ≥80 cm in women; [d]Fat percent (DXA) ≥25% (men) and ≥35% (women); [e]Fasting glucose 100-125 mg/dl (5.6-6.9 mmol/l) (ADA criteria); [f]120-minute glucose 140-199 mg/dl (7.8-11.0 mmol/l) (ADA criteria); [g]Fasting glucose ≥126 mg/dl (7.0 mmol/l) and/or 120-minute glucose ≥200 mg/dl (11.1 mmol/l) (ADA criteria); [h]ln{Insulin(30-minute/fasting)/Glucose(30-minute/fasting)}; [i]10000/√{Glucose fasting x Insulin fasting x mean glucose (F, 30min, 120min) x mean insulin (F, 30min, 120min)} (glucose in mmol/l; insulin in pmol/l); [j](HOMA-S X HOMA-β)/100; [k]Insulinogenic index + ln(Matsuda index). BMI: Body Mass Index; HOMA: Homeostatic Model Assessment models, SD: Standard deviation, LQ: Lower Quadrant, UQ: Upper Quadrant, IUGR: Intra-Uterine Growth Restriction, SGA: Small for Gestation Age.

**Supplemental Table S2a: Lifecourse data in participants with NGT, IFG, IGT and both IFG+IGT (Males)**

| | MALES | | | |
|---|---|---|---|---|
| | **NGT (221)** | **IFG (94)** | **IGT (13)** | **IFG+IGT (24)** |
| | **Median (25th, 75th)** | **Median (25th, 75th)** | **Median (25th, 75th)** | **Median (25th, 75th)** |
| **18 YEARS** | | | | |
| Height (cm) | 170.1[165.7,174.3] | 169.6[166.2,173.4] | 165.6[160.8,168.4] | 169.1[165.6,173.3] |
| BMI (kg/m$^2$) | 18.79[16.97,20.95] | 19.39[18.05,21.88] | 20.25[18.79,22.65] | 20.14[17.58,24.34] |
| Waist circumference (cm) | 70.50[66.20,75.50] | 71.95[67.32,78.35] | 76.55[67.50,82.00] | 75.90[69.05,86.00] |
| Fat (%) | 12.5[8.4,18.3] | 13.3[9.7,20.9] | 22.2[11.6,24.5] | 17.6[10.9,32.1] |
| Fasting plasma glucose (mmol/l) | 5.22[5.05, 5.38] | 5.66[5.55, 5.83] | 5.27[5.11, 5.41] | 5.83[5.60, 6.02] |
| 30 min plasma glucose (mmol/l) | 7.83[7.24, 8.55] | 8.69[7.88, 9.55] | 9.21[7.94,10.04] | 9.32[8.77,10.12 |
| 120 min plasma glucose (mmol/l) | 5.72[5.05,6.44] | 6.30[5.58,7.06] | 8.21[7.94,8.80] | 8.12[7.94,8.91] |
| Fasting plasma insulin (pmol/l) | 48.0[36.0,66.0] | 62.7[46.1,84.9] | 85.8[62.4,113.7] | 70.2[39.7,102.2] |
| 30 min plasma insulin (pmol/l) | 477.0[327.6,705.6] | 466.5[333.6,681.7] | 647.4[330.0,1049.7] | 476.7[386.7,681.0] |
| 120 min plasma insulin (pmol/l) | 258[145.3,372.7] | 280.8[177.0,407.7] | 976.8[567.3,1244.4] | 512.1[380.7,703.5] |
| HOMA ß | 99.00[81.62,119.77] | 100.90[77.72,118.62] | 148.80[122.20,168.70] | 100.30[77.55,144.38] |
| HOMA S | 84.70[61.23,110.50] | 62.15[46.78,84.12] | 47.50[36.90,57.10] | 51.70[38.85,82.60] |
| HOMA DI | 83.76[75.10,98.31] | 64.99[57.62,71.31] | 74.17[61.68,75.50] | 60.87[54.68,67.49] |
| Insulinogenic index | 1.83[1.56,2.15] | 1.57[1.29,1.89] | 1.60[1.30,1.83] | 1.45[1.15,1.75] |
| Matsuda index | 15.44[11.39,21.17] | 12.06[9.34,16.74] | 7.64[5.04,9.54] | 8.51[7.12,12.81] |
| Disposition index | 4.57[4.12,5.01] | 4.11[3.68,4.46] | 3.66[3.02,3.91] | 3.83[3.33,4.17] |

| | | | | |
|---|---|---|---|---|
| **12 YEARS** | | | | |
| Height (cm) | 138.9[132.9,144.8] | 138.1[133.5,143.9] | 131.4[128.8,134.7] | 136.6[133.5,140.9] |
| BMI (kg/m$^2$) | 14.60[13.82,15.66] | 14.86[13.80,15.74] | 13.87[13.28,14.4] | 14.62[13.77,16.49] |
| Waist circumference (cm) | 57.20[54.70,60.20] | 57.50[54.70,61.00] | 55.75[53.38,57.55] | 56.80[54.15,61.15] |
| Fat (%) | 13.4[10.8,17.0] | 14.2[11.4,16.6] | 16.9[12.9,20.7] | 15.7[12.6,20.6] |
| Fasting plasma glucose (mmol/l) | 4.8[4.6,5.1] | 5.0[4.8,5.3] | 4.9[4.7,5.3] | 4.9[4.8,5.3] |
| Fasting plasma insulin (pmol/l) | 27.6[17.8,39.0] | 30.9[18.5,44.1] | 37.2[20.9,45.5] | 26.0[14.3,33.4] |
| HOMA ß | 76.80[56.97,99.07] | 78.20[54.38,96.92] | 93.50[75.8,114.2] | 81.40[44.75,89.65] |
| HOMA S | 148.15[105.62,231.02] | 134.00[91.90,223.32] | 110.30[95.20,179.20] | 158.80[124.50,266.45] |
| HOMA DI | 116.00[100.53,138.83] | 105.94[85.71,134.81] | 103.89[86.45,118.91] | 125.00[105.90,137.79] |
| **6 YEARS** | | | | |
| Height (cm) | 110.0[107.0,113.3] | 110.5[107.8,113.0] | 107.4[106.0,110.1] | 110.0[109.2,112.1] |
| BMI (kg/m$^2$) | 13.45[12.92,14.07] | 13.68[13.11,14.11] | 13.20[13.02,13.26] | 13.51[12.78,13.95] |
| Waist circumference (cm) | 50.1[48.9,51.8] | 50.6[49.2,52.6] | 50.6[47.8,52.3] | 50.6[48.1,52.5] |
| Fat (%) | 17.17[13.53,20.46] | 17.32[14.18,19.91] | 18.90[16.91,20.47] | 19.47[14.86,22.43] |
| Fasting plasma glucose (mmol/l) | 4.9[4.6,5.3] | 5.2[4.8,5.5] | 4.9[4.7,5.2] | 5.1[4.9,5.5] |
| 30 min plasma glucose (mmol/l) | 8.2[6.9,9.2] | 8.1[7.1,9.3] | 7.3[5.9,8.8] | 8.8[6.8,10.6] |
| 120 min plasma glucose (mmol/l) | 5.3[4.5,6.1] | 5.5[4.9,6.4] | 5.2[4.7,6.2] | 5.7[5.1,6.4] |
| Fasting plasma insulin (pmol/l) | 16.8[8.0,26.4] | 21.2[10.2,30.2] | 14.7[7.8,35.3] | 21.1[10.3,33.3] |

| | | | | |
|---|---|---|---|---|
| 30 min plasma insulin (pmol/l) | 138.0[80.4,197.2] | 130.9[92.4,206.9] | 119.7[62.7,148.8] | 148.7[91.3,201.4] |
| 120 min plasma insulin (pmol/l) | 47.8[25.1,76.8] | 53.7[29.6,96.2] | 51.6[28.4,89.4] | 66.5[48.5,84.6] |
| HOMA ß | 52.10[35.08,75.93] | 55.20[36.80,73.85] | 58.30[25.70,78.60] | 53.25[37.12,79.90] |
| HOMA S | 238.75[157.62,468.75] | 191.10[135.80,377.70] | 284.50[124.60,350.30] | 195.30[128.55,389.55] |
| HOMA DI | 135.42[110.69,177.62] | 119.71[93.67,151.00] | 116.60[91.00,202.00] | 107.56[95.94,146.62] |
| Insulinogenic index | 2.85[2.43,3.12] | 2.78[2.48,3.23] | 2.67[2.48,2.99] | 2.80[2.45,3.1] |
| Matsuda index | 58.45[38.62,86.87] | 53.32[32.92,74.97] | 60.26[33.71,95.47] | 46.52[28.95,58.89] |
| Disposition index | 6.80[6.56,7.18] | 6.71[6.32,7.05] | 6.89[6.44,7.05] | 6.45[6.32,6.79] |
| **2 YEARS** | | | | |
| Height (cm) | 82.2[80.6,84.5] | 82.1[80.2,84.3] | 82.0[76.6,82.9] | 81.8[79.8,84.4] |
| Weight (kg) | 9.92[9.25,10.60] | 9.88[9.28,10.40] | 9.70[8.80,10.18] | 9.77[9.28,10.18] |
| **AT BIRTH** | | | | |
| Weight (kg) | 2.7[2.5,2.9] | 2.7[2.5,2.9] | 2.7[2.5,3.0] | 2.6[2.2,2.98] |
| Length (cm) | 48.2[46.6,49.5] | 47.7[46.5,49.05] | 47.4[44.7,48.8] | 48.4[45.9,50.0] |
| Head circumference (cm) | 33.4[32.4,34.2] | 33.1[32.3,34.0] | 33.3[32.2,33.5] | 33.5[31.8,34.1] |

**Supplemental Table S2b: Lifecourse data in participants with NGT, IFG, IGT and both IFG+IGT (Females)**

| | FEMALES | | | |
|---|---|---|---|---|
| | **NGT (213)** | **IFG (23)** | **IGT (23)** | **IFG+IGT (7)** |
| | **Median (25th, 75th)** | **Median (25th, 75th)** | **Median (25th, 75th)** | **Median (25th, 75th)** |
| **18 YEARS** | | | | |
| Height (cm) | 157.2[153.4,161.0] | 157.2[152.4,161.4] | 157.5[155.7,160.5] | 153.6[152.5,157.8] |
| BMI (kg/m$^2$) | 18.08[16.86,20.21] | 17.46[16.89,19.07] | 17.98[16.52,19.76] | 17.29[15.99,18.86] |
| Waist circumference (cm) | 67.45[63.3,71.8] | 65.85[64.5,68.9] | 68.05[62.23,70.32] | 66.65[63.75,71.45] |
| Fat (%) | 28.1[23.35,31.68] | 24.5[21.55,30.49] | 28.91[24.64,33.74] | 24.54[23.2,28.15] |
| Fasting plasma glucose (mg/dl) | 5.1[4.9,5.3] | 5.7[5.6,5.8] | 5.2[4.9,5.3] | 5.7[5.6,6.3] |
| 30 min plasma glucose (mg/dl) | 7.9[7.3,8.7] | 8.9[8.2,9.8] | 9.1[8.6,9.7] | 9.9[8.4,10.8] |
| 120 min plasma glucose (mg/dl) | 6.0[5.4,6.7] | 6.7[6.3,7.3] | 8.3[7.9,9.3] | 8.5[8.2,8.6] |
| Fasting plasma insulin (pmol/l) | 63.0[46.8,80.4] | 76.8[45.3,83.1] | 66.6[55.8,83.4] | 62.4[46.2,81.6] |
| 30 min plasma insulin (pmol/l) | 578.4[40.7.4,774.0] | 510.6[313.5,798.6] | 605.4[385.6,719.4] | 383.8[,246.6,751.2] |
| 120 min plasma insulin (pmol/l) | 360.0[236.5,509.4] | 429.6[350.1,616.2] | 903.0[523.8,1012.2 | 717.0[494.4,1150.2] |
| HOMA ß | 125.5[101.7,143.7] | 110.9[87.1,120.6] | 128.9[117.45,138.75] | 98.1[86.55,112.05] |
| HOMA S | 65.2[51.2,86.75] | 51.6[48,79.4] | 60.6[49.15,68.95] | 63.1[55.95,79.8] |
| HOMA DI | 83.67[74.11,96.7] | 64.53[59.59,67] | 79.38[74.24,86.19] | 63.93[57.72,71.54] |
| Insulinogenic index | 1.77[1.44,2.12] | 1.58[1.25,1.98] | 1.54[1.26,1.65] | 1.18[1.12,1.51] |
| Matsuda index | 12.23[9.36,15.55] | 10.32[7.93,14.16] | 8.41[7.67,10.11] | 10.96[6.89,11.6] |
| Disposition index | 4.27[3.88,4.67] | 4.08[3.51,4.27] | 3.65[3.55,3.77] | 3.56[3.41,3.77] |

| | | | | |
|---|---|---|---|---|
| **12 YEARS** | | | | |
| Height (cm) | 139.6[134.6,146.75] | 140.7[135.05,143.48] | 138.7[133.15,143] | 133.8[129.9,142.95] |
| BMI (kg/m$^2$) | 14.37[13.21,15.97] | 14.25[13.27,15.81] | 14.21[13.29,15.33] | 13.73[12.83,14.6] |
| Waist circumference (cm) | 56[52.72,59.25] | 54.75[51.62,55.9] | 56.05[52.62,58.2] | 54.25[52.12,55.25] |
| Fat (%) | 17.81[14.31,22.85] | 16.44[13.84,18.3] | 18.84[16.5,21.91] | 13.66[11.61,19.16] |
| Fasting plasma glucose (mg/dl) | 4.7[4.4,4.9] | 5.1[4.7,5.3] | 4.9[4.7,5.2] | 5.2[4.8,5.4] |
| Fasting plasma insulin (pmol/l) | 33.9[24.6,47.4] | 35.2[27.8,40.8] | 37.8[24.5,56.9] | 21.0[10.2,40.7] |
| HOMA ß | 96.7[76.95,116.95] | 82.85[72.15,98.75] | 97.5[72.3,121.65] | 52.6[43,83.35] |
| HOMA S | 121.8[88.25,170.15] | 115.25[100.8,141.8] | 109.4[77.6,157.55] | 187.9[122.2,336.85] |
| HOMA DI | 114.5[100.82,136.6] | 100.78[88.35,117.25] | 110.57[87.51,123.78] | 116[95.35,122.21] |
| **6 YEARS** | | | | |
| Height (cm) | 109.4[106.8,112.7] | 107.6[105.2,110.65] | 110[107.6,111.4] | 106.7[106.35,107.1] |
| BMI (kg/m$^2$) | 13.15[12.6,13.79] | 12.89[12.28,13.46] | 12.7[12.31,13.49] | 13.05[12.87,13.15] |
| Waist circumference (cm) | 50[48.5,52.1] | 48.9[47.4,50.55] | 49.9[47.6,50.9] | 49.6[49.05,50.75] |
| Fat (%) | 20.4[17.74,24.07] | 19.73[17.2,22.62] | 22[19.42,23.48] | 16.63[14.14,18.52] |
| Fasting plasma glucose (mg/dl) | 4.8[4.6,5.2] | 5.1[4.8,5.4] | 4.8[4.4,4.9] | 5.3[4.5,5.3] |
| 30 min plasma glucose (mg/dl) | 8.1[7.2,9.1] | 9.2[7.9,10.3] | 8.1[7.3,9.5] | 8.7[7.2,9.1] |
| 120 min plasma glucose (mg/dl) | 5.6[4.9,6.2] | 5.3[4.9,6.7] | 6.0[5.3,6.8] | 5.1[4.3,5.5] |
| Fasting plasma insulin (pmol/l) | 20.0[9.9,31.3] | 16.2[9.0,20.9] | 21.8[8.40,37.5] | 19.6[8.3,29.3] |

| | | | | |
|---|---|---|---|---|
| 30 min plasma insulin (pmol/l) | 163.5[109.9,202.8] | 176.2[108.8,264.0] | 138.1[115.3,213.5] | 132.5[98.2,202.0] |
| 120 min plasma insulin (pmol/l) | 66.1[39.5,97.9] | 45.0[22.7,104.9] | 119.5[47.6,172.3] | 33.3[30.1,61.6] |
| HOMA ß | 65.3[44.75,89.1] | 47.4[36.27,72.52] | 72.25[46.83,85.57] | 48.1[29.3,91.7] |
| HOMA S | 199.35[130.7,388.1] | 242.25[197.25,419.97] | 192.3[115.05,361.2] | 200.3[151.35,464.15] |
| HOMA DI | 132.33[102.11,175.87] | 118.67[94.24,144.88] | 129.25[98.25,167.65] | 134[114.1,146.5] |
| Insulinogenic index | 2.96[2.67,3.3] | 2.91[2.65,3.36] | 2.77[2.61,3.22] | 2.93[2.44,3.19] |
| Matsuda index | 46.8[32.78,67.31] | 50.13[38.84,61.82] | 53.53[25.41,72.58] | 43.86[38.25,77.93] |
| Disposition index | 6.8[6.47,7.11] | 6.76[6.56,7.06] | 6.72[6.27,6.94] | 6.77[6.71,6.92] |
| **2 YEARS** | | | | |
| Height (cm) | 81.3[78.7,83.1] | 80.2[77.8,81.4] | 79.6[78.5,82.7] | 78.9[78.1,80.7] |
| Weight (kg) | 9.22[8.61,10.07] | 8.69[8.05,9.54] | 9.04[8.33,10.01] | 8.64[8.41,9.38] |
| **AT BIRTH** | | | | |
| Weight (kg) | 2.55[2.34,2.8] | 2.3[2.16,2.5] | 2.75[2.5,2.9] | 2.5[2.08,2.7] |
| Length (cm) | 47.4[46.3,48.5] | 46[45.2,47.95] | 47.5[45.45,48.15] | 45.8[44.95,46.5] |
| Head circumference (cm) | 32.7[31.9,33.5] | 32.2[31.35,33.1] | 33.1[31.8,33.7] | 32.2[30.75,32.5] |

**Supplemental Table S3: Associations of lifecourse measurements with IFG and IGT, each compared with NGT, at 18 years**

| | Males | | | | Females | | | |
|---|---|---|---|---|---|---|---|---|
| | IFG (94) | | IGT (13) | | IFG (23) | | IGT (23) | |
| | Beta | P-value | Beta | P-value | Beta | P-value | Beta | P-value |
| **18 YEARS** | | | | | | | | |
| Height (cm) | -0.02 | 0.711 | -0.075 | 0.17 | -0.058 | 0.353 | -0.002 | 0.972 |
| BMI (kg/m$^2$) | 0.102 | 0.061 | 0.096 | 0.079 | -0.006 | 0.927 | -0.038 | 0.537 |
| Fat (%) | 0.109 | 0.043 | 0.164 | 0.003 | -0.053 | 0.396 | 0.049 | 0.437 |
| Fasting plasma glucose (mg/dl) | 0.729 | <0.000 | 0.095 | 0.009 | 0.614 | <0.000 | 0.068 | 0.163 |
| 30 min plasma glucose (mg/dl) | 0.314 | <0.000 | 0.208 | <0.000 | 0.224 | <0.000 | 0.255 | <0.000 |
| 120 min plasma glucose (mg/dl) | 0.205 | <0.000 | 0.509 | <0.000 | 0.143 | 0.004 | 0.563 | <0.000 |
| Fasting plasma insulin (pmol/l) | 0.188 | <0.000 | 0.137 | 0.011 | 0.029 | 0.64 | 0.086 | 0.168 |
| 30 min plasma insulin (pmol/l) | -0.013 | 0.812 | 0.078 | 0.156 | -0.093 | 0.136 | -0.023 | 0.719 |
| 120 min plasma insulin (pmol/l) | 0.037 | 0.459 | 0.398 | <0.000 | 0.08 | 0.173 | 0.34 | <0.000 |
| HOMA ß | -0.05 | 0.366 | 0.151 | 0.006 | -0.225 | <0.000 | 0.055 | 0.371 |
| HOMA S | -0.235 | <0.000 | -0.175 | 0.001 | -0.058 | 0.354 | -0.089 | 0.153 |
| HOMA DI | -0.548 | <0.000 | -0.153 | 0.001 | -0.418 | <0.000 | -0.108 | 0.064 |
| Insulinogenic index | -0.194 | <0.000 | -0.084 | 0.119 | -0.094 | 0.128 | -0.142 | 0.022 |
| Matsuda index | -0.199 | <0.000 | -0.262 | <0.000 | -0.103 | 0.09 | -0.235 | <0.000 |
| Disposition index | -0.176 | 0.001 | -0.177 | 0.001 | -0.143 | 0.017 | -0.255 | <0.000 |

| | | | | | | | | |
|---|---|---|---|---|---|---|---|---|
| **12 YEARS** | | | | | | | | |
| Height (cm) | -0.01 | 0.856 | -0.134 | 0.015 | -0.045 | 0.472 | -0.075 | 0.233 |
| BMI (kg/m$^2$) | 0.064 | 0.249 | -0.058 | 0.294 | -0.007 | 0.915 | -0.049 | 0.435 |
| Fat (%) | 0.029 | 0.608 | 0.126 | 0.028 | -0.061 | 0.342 | 0.022 | 0.735 |
| Fasting plasma glucose (mg/dl) | 0.206 | <0.000 | -0.027 | 0.627 | 0.247 | <0.000 | 0.201 | 0.001 |
| Fasting plasma insulin (pmol/l) | 0.003 | 0.951 | -0.014 | 0.807 | -0.067 | 0.286 | -0.004 | 0.944 |
| HOMA ß | -0.069 | 0.215 | -0.005 | 0.926 | -0.165 | 0.008 | -0.084 | 0.172 |
| HOMA S | -0.011 | 0.838 | 0.016 | 0.776 | 0.049 | 0.435 | -0.008 | 0.901 |
| HOMA DI | -0.126 | 0.023 | 0.018 | 0.744 | -0.129 | 0.037 | -0.132 | 0.032 |
| **6 YEARS** | | | | | | | | |
| Height (cm) | 0.036 | 0.519 | -0.034 | 0.536 | -0.137 | 0.027 | -0.026 | 0.674 |
| BMI (kg/m$^2$) | 0.108 | 0.05 | -0.089 | 0.106 | -0.073 | 0.24 | -0.072 | 0.243 |
| Fat (%) | 0.012 | 0.823 | 0.116 | 0.036 | -0.106 | 0.088 | 0.046 | 0.456 |
| Fasting plasma glucose (mg/dl) | 0.184 | <0.000 | 0.056 | 0.276 | 0.185 | 0.003 | -0.007 | 0.114 |
| 30 min plasma glucose (mg/dl) | 0.066 | 0.235 | -0.005 | 0.932 | 0.151 | 0.015 | -0.022 | 0.728 |
| 120 min plasma glucose (mg/dl) | 0.088 | 0.11 | 0.057 | 0.301 | -0.012 | 0.841 | 0.111 | 0.076 |
| Fasting plasma insulin (pmol/l) | 0.114 | 0.038 | 0.03 | 0.579 | -0.057 | 0.361 | -0.004 | 0.954 |
| 30 min plasma insulin (pmol/l) | 0.079 | 0.149 | -0.081 | 0.139 | 0.066 | 0.293 | -0.039 | 0.534 |
| 120 min plasma insulin (pmol/l) | 0.072 | 0.188 | 0.056 | 0.309 | -0.15 | 0.015 | 0.114 | 0.064 |
| HOMA ß | 0.026 | 0.645 | 0.001 | 0.99 | -0.122 | 0.052 | 0.008 | 0.133 |

| | | | | | | | | |
|---|---|---|---|---|---|---|---|---|
| HOMA S | -0.111 | 0.046 | -0.032 | 0.564 | 0.042 | 0.508 | -0.006 | 0.921 |
| HOMA DI | -0.18 | 0.001 | -0.067 | 0.219 | -0.073 | 0.245 | -0.016 | 0.804 |
| Insulinogenic index | 0.083 | 0.134 | -0.079 | 0.155 | 0.026 | 0.676 | -0.014 | 0.823 |
| Matsuda index | -0.154 | 0.005 | -0.013 | 0.816 | -0.001 | 0.993 | 0.024 | 0.699 |
| Disposition index | -0.107 | 0.051 | -0.11 | 0.046 | 0.037 | 0.549 | -0.074 | 0.238 |
| **2 YEARS** | | | | | | | | |
| Height (cm) | -0.003 | 0.950 | -0.083 | 0.132 | -0.097 | 0.116 | -0.088 | 0.154 |
| Weight (kg) | 0.009 | 0.874 | -0.070 | 0.204 | -0.153 | 0.013 | -0.057 | 0.357 |
| **AT BIRTH** | | | | | | | | |
| Weight (kg) | -0.033 | 0.563 | -0.048 | 0.399 | -0.237 | <0.000 | 0.081 | 0.196 |
| Length (cm) | -0.021 | 0.708 | -0.044 | 0.425 | -0.155 | 0.012 | -0.067 | 0.267 |
| Head circumference (cm) | -0.097 | 0.08 | -0.035 | 0.525 | -0.148 | 0.017 | 0.022 | 0.722 |

Beta and p-values are generated using linear regression model with IFG and IGT as separate exposures, models are separately analysed for sex.

**Supplemental Table S4: Methods for anthropometry and body composition measurements in children**

| **Anthropometry** | | | |
|---|---|---|---|
| Measurement | Time point | Method | Least count |
| Weight | Within 72 hours of birth | Salter spring balance (Salter Abbey, Suffolk, U.K.) | Nearest 50 g |
| | 6 monthly intervals up to 18 years | Electronic weighing scales (ATCO Healthcare Ltd, Mumbai, India) | Nearest 10 g |
| Crown heel length (Supine) | Within 72 hours of birth and 6 monthly upto 2 years | Portable Pedobaby Babymeter (ETS J.M.B., Brussels, Belgium) | Nearest 0.1 cm |
| Standing height | 6 monthly intervals from 2- 5 years | Portable Harpenden stadiometer (Microtoise, CMS Instruments Ltd, London, UK) | Nearest 0.1 cm |
| | 6 monthly intervals from 5-18 years | Wall-mounted stadiometer (Microtoise, CMS Instruments Ltd, London, UK) | Nearest 0.1 cm |
| **Body composition by Dual Energy X-ray Absorptiometry (DXA)** | | | |
| Total fat and lean mass; % body fat | 6, 12 year follow up in children | Lunar DPX-IQ 240, Lunar Corporation, Madison, USA | NA |
| | 18 year follow up | Lunar Prodigy, GE Healthcare, Madison, USA | NA |

**Supplemental Table S5**: **Quality assessments for glucose and insulin measurements in the Pune Maternal Nutrition Study (1993- 2013).**

| | **1994-97** | **2000-02** | **2006-08** | **2013-14** |
|---|---|---|---|---|
| **Glucose measurement** | | | | |
| Method | GOD-POD | GOD-POD | GOD-POD | GOD-POD |
| Equipment | Spectrum; Abbott, Irving, TX | Spectrum; Abbott, Irving, TX | Alcyon; Abbott, Irving, TX | Hitachi 902, Roche Diagnostics GmbH, Germany |
| Centrifugation temperature | Room temperature | Room temperature | 4°C | 4°C |
| Processed on | Same day | Same day | Same day | Same day |
| Internal QC | Yes | Yes | Yes | Yes |
| Internal CV (%) | <3 | <3 | <3 | <3 |
| External QC | NA | NA | BioRad EQAS | BioRad EQAS |
| EQAS CV (%) | NA | NA | 3.4 | 2.7 |
| **Insulin measurements** | | | | |
| Method | 1-step chemiluminescent immunoenzymatic assay* | 2 site fluoroimmunometric assay | 2 site fluoroimmunometric assay | ELISA |
| Equipment | Access Immunoassay System (Sanofi Pasteur Diagnostics, Chaska, Minn) | Delfia technique | Delfia technique | Mercodia AB, SE-754 50 Uppsala, Sweden |
| Centrifugation temperature | Room temperature | Room temperature | 4°C | 4°C |

| | | | | |
|---|---|---|---|---|
| Sensitivity | 2.3 pM/L | 3 pM/L | 3 pM/L | 6 pM/L |
| Calibrated against | WHO 1st IRP (66/304) | WHO 1st IRP (66/304) | WHO 1st IRP (66/304) | WHO 1st IRP (66/304) |
| Internal QC | Yes | Yes | Yes | Yes |
| Internal CV | NA | NA | 7.7%. | 6.7% |
| External QC | No | UKEQAS | UKEQAS | UKEQAS |
| External CV | NA | <20 pM/L: 8.9% 45-90 pM/L: 6.8% | <20 pM/L: 8.2% 20-45 pM/L: 6.9% >45 pM/L: 8.6% | <20 pM/L: 6.4% 20-45 pM/L: 7.6% >45 pM/L: 5.5% |

GOD POD: Glucose oxidase peroxidase; QC: Quality control, CV: Coefficient of variation

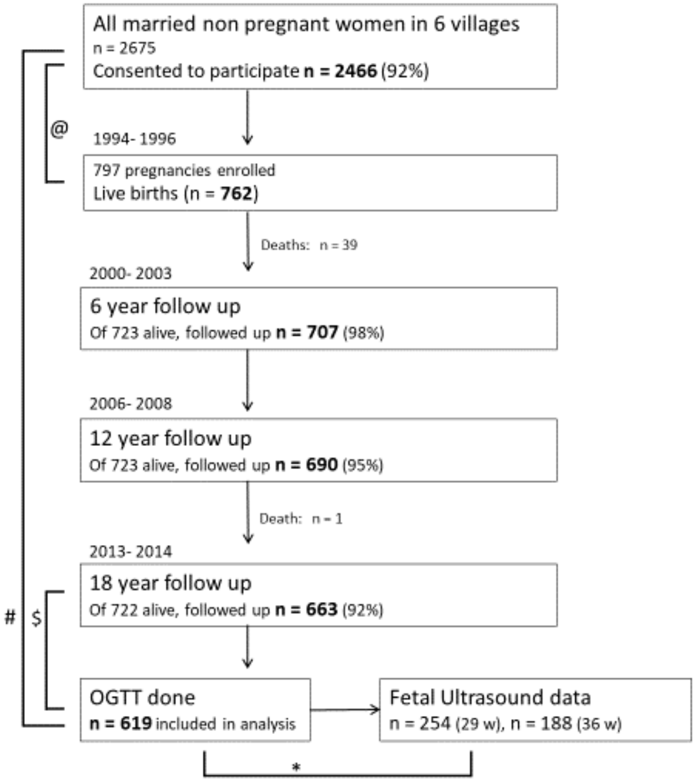

**Supplemental figure S1**: STROBE flow diagram for the Pune Maternal Nutrition Study.

Legend:

@: The 797 pregnant F0 women included in the PMNS had similar education and height but were younger (20y vs 22y) and thinner (18.1 vs 18.4 kg/m2) compared to those excluded.

#: The mothers of the 619 participants who had an OGTT at 18 years were similar in height and education at baseline to the 1,847 of 2,466 women who did not become pregnant during the initial study and hence did not get enrolled in the PMNS, $p > 0.05$ for both.

$: The 619 participants who had an OGTT at 18 years were similar to the 103 who did not have an OGTT, in birth length and birth weight, $p > 0.05$ for both. Their parents were similar in socioeconomic status (SLI Median 27 in both, $p > 0.05$). OGTT: Oral Glucose Tolerance Test.

**Supplemental figure S2: Comparative sizes across lifecourse of participants with glucose intolerance**

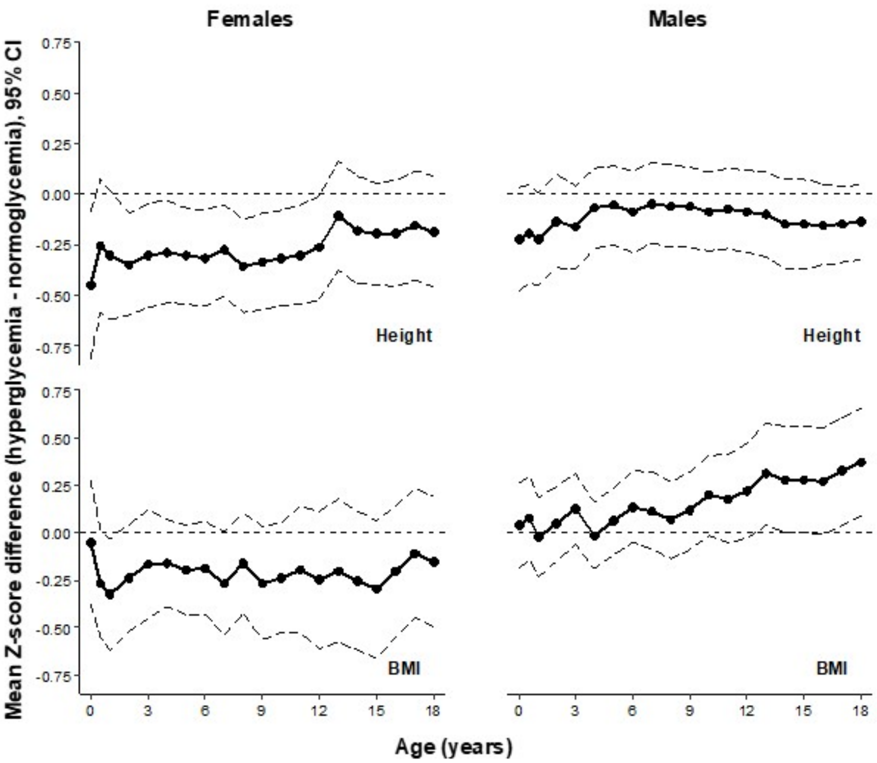

Plots show comparative size (height and BMI expressed as Z scores) of men and women with glucose intolerance at age 18 years relative to those with normal glucose tolerance (represented by the 0 line). Z scores were derived using the WHO growth reference. The dark line represents the median and the dashed lines represent 95% confidence intervals.

**Supplemental figure S3:** ROC curves showing the prediction of glucose intolerance at 18 years (outcome) by 6-year (5a) and 12-year (5b) fasting plasma glucose, and other predictors, in the PMNS cohort

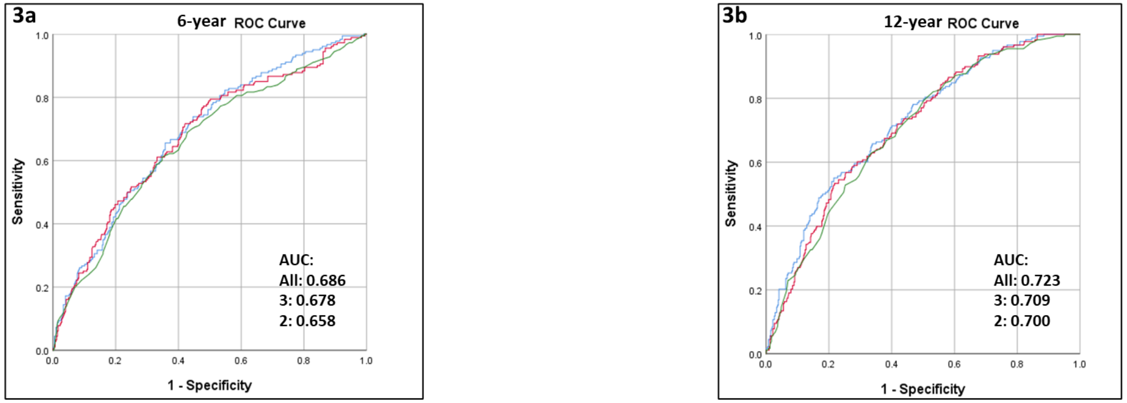

All predictors: Parental glucose intolerance, underweight & overweight/obesity, birth length, fasting glucose at 6y, child's SLI, height & DXA fat % at 18y, and sex
3 predictors: Birth length, , fasting glucose at 6y and sex
2 predictors: fasting glucose at 6y and sex

All predictors: Parental glucose intolerance, underweight & overweight/obesity, birth length, fasting glucose at 12y, child's SLI, height & DXA fat % at 18y, and sex
3 predictors: Birth length, , fasting glucose at 12y and sex
2 predictors: fasting glucose at 12y and sex

**Supplemental figure S4: Glucose intolerance in the PCS and Extended PMNS cohorts**

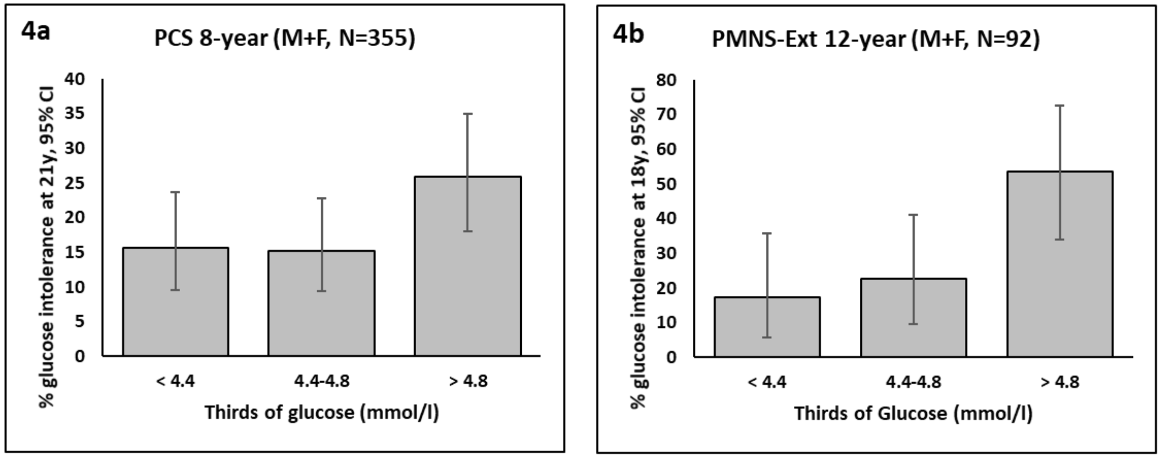

Figures 4a and 4b show the prevalence of glucose intolerance at 21y in the PCS and at 18y in the extended PMNS cohort according to thirds of fasting plasma glucose at 8y and 12y respectively.
